# Deep Learning Analysis of Figure Copying Tasks for Parkinson’s Disease Detection with GAN-Based Data Augmentation

**DOI:** 10.1101/2025.04.15.25325847

**Authors:** Marta Vallejo, Jane Alty, Stephen L. Smith

**Affiliations:** School of Computer Sciences, Heriot-Watt University, Edinburgh Campus, Edinburgh, EH14 4AS, Scotland, UK; School of Medicine, University of Tasmania, Churchill Ave, Hobart, 7000, Tasmania, Australia; Wicking Dementia Research and Education Centre, University of Tasmania, College Road, Hobart, 7000, Tasmania, Australia; Royal Hobart Hospital, 48 Liverpool Street, Hobart, 7000, Tasmania, Australia; School of Physics, Engineering and Technology, University of York, Heslington, York, YO10 5DDS, England, UK

**Keywords:** Bradykinesia, Deep Learning, Data Augmentation, Generative Adversarial Networks, Geometric Transformations, Drawing Task Analysis

## Abstract

Early and accurate diagnosis of Parkinson’s disease (PD) is essential for enabling timely treatment and effective disease management. In this study, we propose a deep learning approach to automate PD detection using convolutional neural networks (CNNs) trained on images derived from spiral drawing tasks performed by patients and healthy controls. These drawings were collected using a digital pen and tablet, which captured dynamic signals during the task. A total of 82 drawings from 56 PD patients and 26 control subjects were converted into three visual modalities: time, pressure, and pen angle relative to the X and Y axes. Given the small dataset size, we implemented several data augmentation strategies to increase training diversity and balance class distributions. These included traditional geometric transformations, as well as synthetic augmentation using generative adversarial networks (GANs) and deep convolutional GANs (DCGANs). All augmented datasets were used to train a CNN classifier.

Among all experiments, the highest classification performance was achieved using representations derived from pen pressure, combined with traditional augmentation techniques, reaching 80.14% accuracy and a Kappa value of 0.57. This modality consistently outperformed both time and angle-based representations. While GAN and DCGAN models produced visually varied images, they required extensive training epochs to generate sufficient sample diversity, limiting their current practicality. These findings demonstrate the potential of combining CNNs with drawing-based representations and augmentation methods to create a non-invasive, rapid screening tool for PD. Future work will aim to expand the dataset, investigate more advanced model architectures such as transformers and attention-based networks, and further explore the impact of input resolution and colour mapping on performance.

## 1. Introduction

Parkinson’s disease (PD) is a progressive neurodegenerative disorder that significantly impacts daily functioning and quality of life. There is strong evidence that early detection and treatment of PD, with medications that increase dopamine in the brain, improves motor symptoms and quality of life (de Bie et al., 2020).

Early and accurate detection of PD can be challenging, as motor characteristics may be subtle and gradually occur, often dismissed as physiological ageing (Bloem et al., 2021). An objective test that is rapid, low-cost, and easy to administer would be valuable to aid in the early detection of PD. To address this need, several machine-based systems have been developed to identify early features of PD using input modalities such as voice recordings, digital drawings, and handwriting samples. Pereira et al. (2015) proposed using Deep Learning (DL) techniques and drawing data to support the diagnosis of PD. The authors also designed and uploaded a dataset called “HandPD”, with images collected from the handwriting exams. More and more researchers are applying DL techniques in this research context. In the published work of (Alissa et al., 2022), a relatively simple Convolutional Neural Network (CNN) classifier trained with pressure representation images of pentagon drawings is used to discriminate PD from healthy controls.

Although DL is a remarkable tool for handling many computer vision tasks, it relies heavily on extensive data to avoid over-fitting. Unfortunately, it is challenging in the medical field to access such large-size samples (Chlap et al., 2021). To address this issue, data augmentation has become a popular method for increasing the size of a training dataset, which can also enhance the invariance and provide robustness for training DL models (Shorten and Khoshgoftaar, 2019).

In this work, participants were asked to draw pentagon figures using a digital pen and tablet, which recorded detailed information about their drawing movements. The resulting time-series signals were then extracted and converted into image-based data representations. Three data representations are introduced: pressure, time, and angle. Using data augmentation techniques, the generated training set is loaded into the built CNN model to train a classifier to distinguish PD patients and controls (Fig. 1).

**Figure 1:**
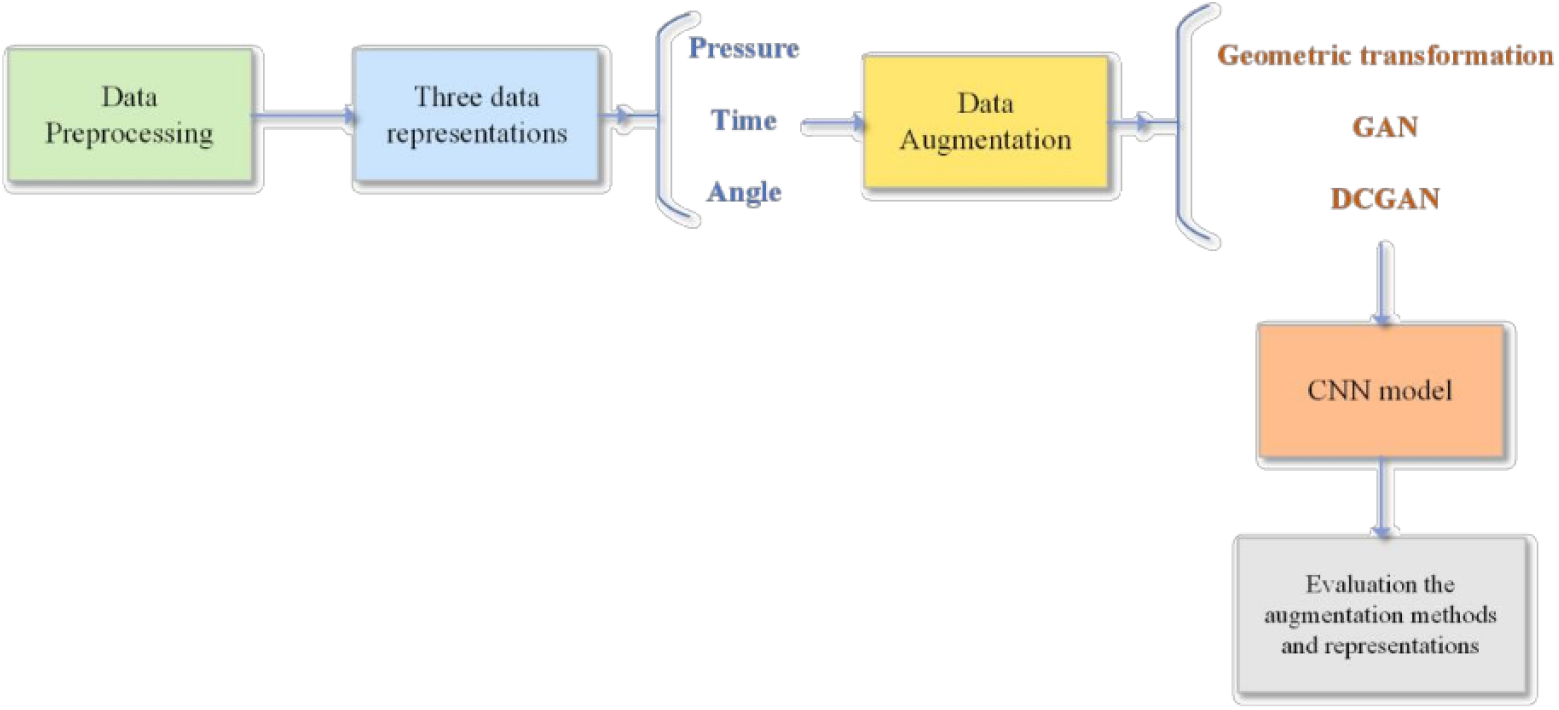
Flow char of the methodology followed.

In this study, we explore and compare various data augmentation techniques, including geometric transformations and generative adversarial networks (GANs), to address the challenges posed by limited dataset size. Using a dataset of pentagon drawings, the primary goal is to evaluate the classification performance of a CNN model under different configurations. Specifically, this work focuses on two sub-objectives: first, to compare different data representations and identify the most informative option for the predictive model, and second, to assess the impact of various augmentation techniques on classification performance and its stability.

The remainder of this paper is organised as follows. Section 2 presents a theoretical background concerning PD and the data augmentation techniques employed in this work. Section 3 outlines the dataset and the data preprocessing and describes the experiments performed. Section 4 presents the experimental results and their analysis. Section 5 discusses our work, comments on the experimental results in detail, and lists some limitations. Section 6 briefly concludes the paper and highlights future work directions.

## 2. Background

### 2.1 Parkinson’s Disease

PD is the second-most common neurodegenerative disorder, affecting approximately 2–3% of individuals over the age of 65 (Tanner and Goldman, 1996). According to global estimates, approximately 10 million people world-wide are affected by PD, with about one million cases in the United States, 1.2 million in Europe, and a projected two million cases in China by 2030 (Dorsey et al., 2018). For all PD patients, treatment is symptomatic, focused on improvement in motor (e.g., tremor, rigidity, bradykinesia) and non-motor (e.g., constipation, cognition, mood, sleep) signs and symptoms. No disease-modifying pharmacologic treatments are available (Armstrong and Okun, 2020). Nevertheless, early diagnosis and treatment of PD are crucial, as they can help manage symptoms more effectively and improve patients’ quality of life (Schrag et al., 2015).

However, an accurate early diagnosis can be challenging because the symptoms may overlap with other conditions (Bloem et al., 2021). PD diagnosis is principally clinical, based on identifying some combination of the cardinal motor signs of bradykinesia, rigidity, tremor, and postural instability (Gelb et al., 1999). Additionally, diagnostic imaging techniques that support clinical diagnosis, such as dopamine transporter scans, are significantly costly, non-specific and inaccessible to many around the world. Therefore, there is still a need for rapid and cost-effective screening tests that can provide objective results to support the detection of PD at an early stage.

Micrographia, characterised by an abnormal reduction in writing size, is one of the most commonly reported and easily detectable handwriting abnormalities in patients with PD, affecting approximately 63% of individuals with the condition (Letanneux et al., 2014). Studies have also shown that kinematic aspects of handwriting—such as size, speed, acceleration, and stroke length—are impaired in PD from its early stages (Tucha et al., 2006). Consequently, handwriting analysis may be a useful method for the early detection of the disease. These motor deficits can be observed through standardised handwriting tasks, which may contribute to the early identification of PD-related motor impairment.

### 2.2 Augmentation Techniques for Deep Learning

DL methods, and in particular CNNs, have led to an enormous break-through in a wide range of computer vision tasks, primarily using large-scale annotated datasets (Frid-Adar et al., 2018). However, medical imaging datasets are often limited and imbalanced, meaning that certain classes are underrepresented, which can lead to biased learning and overfitting when training deep learning models. Data imbalance occurs when the distribution of samples across classes is uneven, resulting in models that may perform well on the majority class but poorly on minority classes. Overfitting, in this context, can lead to a model that performs perfectly on the training dataset but fails to achieve high accuracy on new testing datasets (Elgendi et al., 2021).

Data augmentation encompasses a suite of techniques that enhance the size and quality of training datasets so that better DL models can be built using them. The image augmentation algorithms include traditional geometric transformations, colour space augmentations, kernel filters, mixing images, random erasing, feature space augmentation, adversarial training, GANs, neural style transfer, and meta-learning (Shorten and Khoshgoftaar, 2019).

#### 2.2.1 Traditional Geometric Transformations

The earliest demonstrations showing the effectiveness of data augmentation come from simple traditional transformations such as horizontal flipping, colour space augmentations, geometric rotation and random cropping. When applying geometric transformation methods, attention should be paid to the “security” of their strategies. The safety of the data augmentation method refers to its likelihood of preserving the label post-transformation. For example, rotations and flips are generally safe on ImageNet challenges, such as cats versus dogs, but not for digit recognition tasks like six versus nine (Shorten and Khoshgoftaar, 2019).

The GAN architecture first proposed by Goodfellow et al. (2020) is a framework for generative modelling through adversarial training (Fig. 2). The approach for understanding GANs is the analogy of a cop and a counterfeiter. The counterfeiter (generator network) takes in some form of input, like a random vector, image, or text. The counterfeiter learns to produce an output such that the cop (discriminator network) cannot tell if the output is real or fake (created by the generator network). The impressive performance of GANs has led to a growing interest in applying them to medical data augmentation Chen et al. (2022). These networks can generate new training data that results in better-performing classification models. For example, in 2018, Frid-Adar et al. (2018) used this model for liver lesion classification. The GAN-based data augmentation improved the classification performance from 78.6% sensitivity and 88.4% specificity using classic augmentations to 85.7% sensitivity and 92.4% specificity using GAN-based data augmentation.

**Figure 2:**
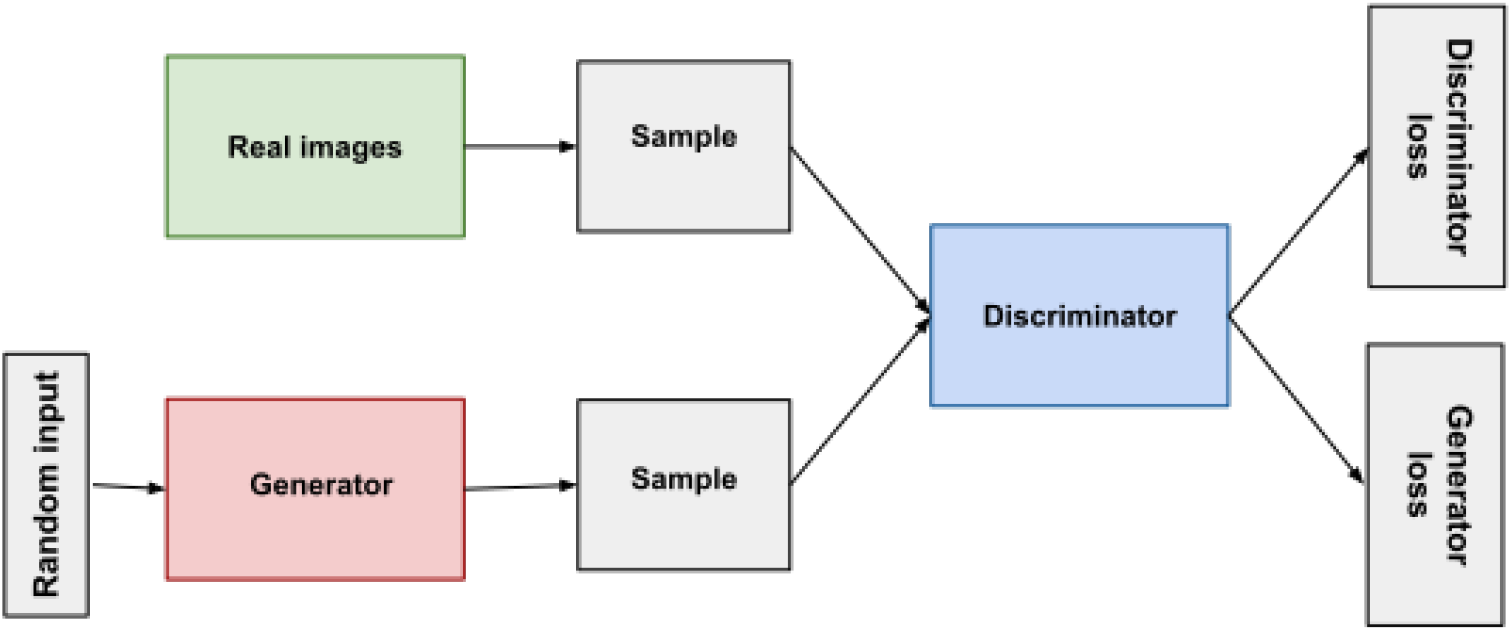
General structure of a generative adversarial network.

#### 2.2.2 Deep convolutional generative adversarial network for data augmentation

Many new architectures have been proposed to extend the concept of GAN. Among them, DCGANs, progressively growing GANs, CycleGANs, and conditional GANs showed great potential for applications for data augmentation (Shorten and Khoshgoftaar, 2019). The DCGAN (Radford et al., 2015) architecture was proposed to expand on the internal complexity of the generator and discriminator networks. The DCGAN architecture presents a strategy for using convolutional layers in the GAN framework to produce higher-resolution images instead of multilayer perceptrons (Fig. 3).

**Figure 3:**
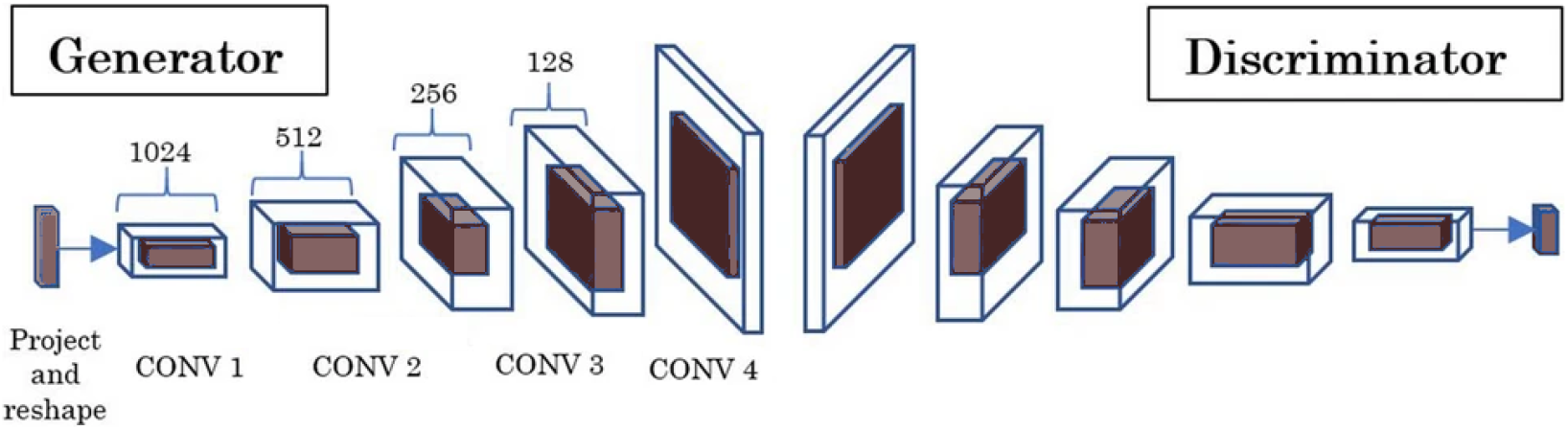
General structure of a deep convolutional generative adversarial network.

The idea behind DCGAN was to increase the complexity of the generator network to project the input into a high-dimensional tensor and then add deconvolutional layers to go from the projected tensor to an output image. These deconvolutional layers will expand on the spatial dimensions, for example, from (14 *×* 14 *×* 6) to (28 *×* 28 *×* 1), whereas a convolutional layer will decrease the spatial dimensions from (14 *×* 14 *×* 32) to (7 *×* 7 *×* 64).

The DCGAN was successfully tested on the LSUN interior bedroom image dataset ^1^, with images of (64 *×* 64 *×* 3), for a total of 12,288 pixels.

## 3. Methodology

### 3.1 Data Acquisition

For this study, data were collected by clinicians at Leeds Teaching Hospitals NHS Trust UK. The dataset comprises information from 87 participants, including 58 patients diagnosed with PD and 29 age-matched healthy controls. Patients were recruited from neurology clinics and diagnosed by specialist consultants according to the Queen Square Brain Bank Criteria (Gibb and Lees, 1989). Healthy controls were recruited as the spouses or friends of patients and were eligible for inclusion if they had no history of neurological disorders. All procedures were conducted in accordance with the guidelines of the relevant institutional review board, ensuring ethical standards were met throughout the study. The study was approved by the corresponding ethics review board. Written informed consent was obtained from every participant prior to data collection.

For the drawing tasks, each subject performed two drawings with each hand. All subjects were asked to draw an Archimedean spiral pentagon on top of a template image using an inking stylus and a digitising, pressure-sensitive Wacom tablet (Wacom Technology Corporation) measuring 20.3 cm *×* 32.5 cm. Participants were instructed to draw the figures as accurately and as fast as possible. The tablet recorded data with a constant sample ratio of 200 Hz. There were no specific rules on how participants should follow the template; some started drawing from the inside of the pentagon, while others began from the outside. This variation is particularly relevant, as decrement, characterised by a gradual loss of pressure towards the end of the task, is a significant feature of PD. The datasets include information from 87 subjects, comprising 58 patients and 29 age-matched healthy controls. Detailed demographic and clinical characteristics of the participants are presented in Table 1.

**Table 1:** Demographic and clinical characteristics of the study participants. Values are presented as mean (standard deviation, range) where applicable. MDS-UPDRS: Movement Disorder Society – Unified Parkinson’s Disease Rating Scale; MoCA: Montreal Cognitive Assessment; LEDD: Levodopa Equivalent Daily Dose; M: male; F: female; R: right-handed; L: left-handed.

| Characteristics | PD group (n = 58) | Controls (n = 29) |
| --- | --- | --- |
| Age, years | 69.2 (8.4, 44–85) | 66.1 (7.6, 50–79) |
| Gender, M:F | 38:20 | 5:24 |
| Handedness, R:L | 51:7 | 22:7 |
| Disease duration, years | 6.2 (4.7, 0.5–20) | - |
| MDS-UPDRS Part 3 | 28.8 (11.5, 3–56) | 1.9 (2.3, 0–8) |
| MoCA score | 23.1 (4.1) | 26.3 (3.0) |
| LEDD, mg/day | 662.7 (560.9) | - |

During the individual drawing test, specific information is extracted and stored as time series data to evaluate the movements performed. Each multivariate sample contains information corresponding to the timestamp, including coordinates X and Y of each pen position, the angles of the pen relative to the X and Y planes, and the relative pressure applied to the tablet. Roughly speaking, each test comprises n rows (the end time of the test) and six columns, which stand for the aforementioned six extracted signals.

These extracted signals have a range of values. The timestamp entries are monotonic integer values starting from 0. Coordinates X and Y and pressure values are represented in the range of [− 0, 1], and the angle of the pen is in the range [−1, 1]. In particular, it can be interpreted that when zero pressure value is collected, the pen at this position is not in contact with the tablet. The labelled data is also limited and imbalanced, so data augmentation techniques, such as geometric transformation methods and GAN models, generate synthetic medical images to extend the datasets. This can enhance the invariance and provide robustness for training models like CNNs.

### 3.2 Data Pre-processing

#### 3.2.1 The Generation of Images

The problem of distinguishing PD patients from control individuals can be modelled as an image recognition task. Using this approach, our CNN model is expected to learn distinctive features in the drawing tasks that differentiate PD patients from healthy individuals based on subtle variations in movement patterns.

To convert the information collected by the pen into an image, for each subject experiment, the coordinates X and Y corresponding to each times-tamp were extracted and displayed as the trajectory described by the pen. In this way, two-dimensional black-and-white images can be obtained. Fig. 4 illustrates the converted images of control and patient subjects in terms of the pentagon dataset.

**Figure 4:**
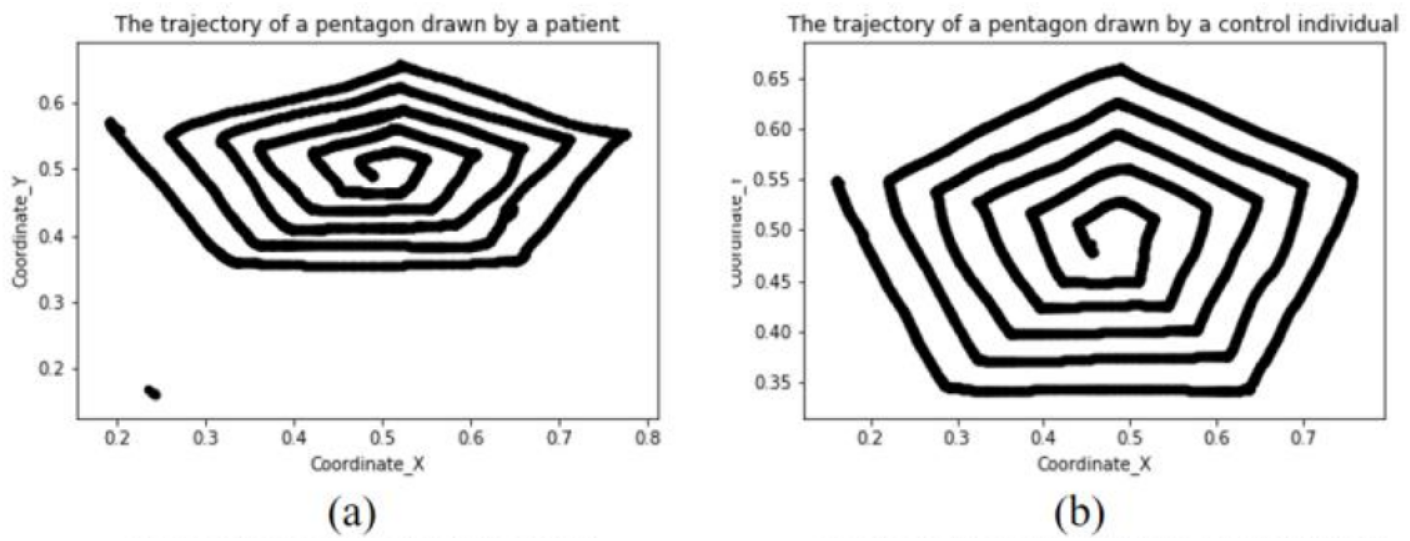
(a) The trajectory of a pentagon drawn by a PD patient and (b) the trajectory of a pentagon drawn by a control individual.

#### 3.2.2 Data Representation - Colour

This work proposes different representations of images to find which representation best fits our CNN model. Specifically, the three data representations are pressure representation, time representation and angle representation. To achieve this, the representation data columns corresponding to each timestamp are extracted, and then the value is mapped to the corresponding colour using a colour map. Then, the final image will be loaded into the CNN model as input. The output performance of these three representations will also be compared and discussed in the results section.

Each representation value can be normalised and then mapped to the corresponding colour according to the chosen colour map. This work focuses on using a coloured instead of a grayscale colour map to implement the data representation, as colour images are expected to give better results. Specifically, a perceptually uniform colour map is more appropriate since the represented information is sequential. The Viridis and greys colour maps, as displayed in Fig. 5, were chosen for colour mapping. The main difference is that the values of the image are stored in three channels (RGB) in the case of the Viridis colour map, while the grey colour map only uses one channel (Sisneros et al., 2016).

**Figure 5:**
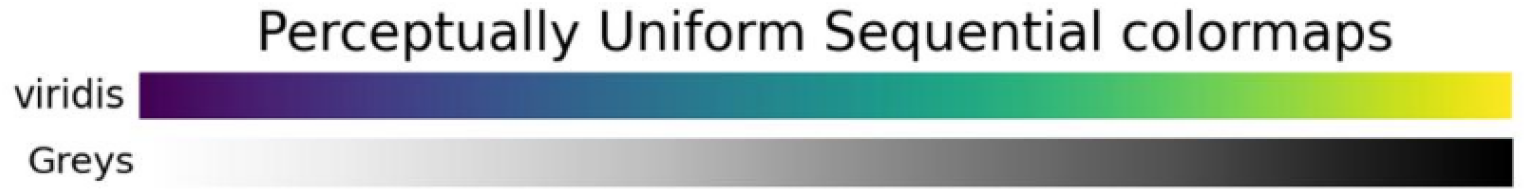
The Viridis and the grey colour maps.

#### 3.2.3 Data Representation - Pressure

As for pressure representation, it has been found that pressure decreases with the progression of PD (Bu et al., 2013). If information on pressure magnitude can be included in the trajectory of drawings, then the pressure representation images can be used to discriminate between patients and controls. Specifically, the trained classifier can obtain the features of the pressure magnitude and distinguish controls from patients. Fig. 6 shows the pressure representation images of the pentagon dataset for both the patient and control groups. The colours of the patient group can be intuitively seen to be significantly darker than those of the control group. According to the Viridis colour map, the darker the colour is, the smaller the value. Therefore, this phenomenon can be explained by the fact that PD patients tend to draw with less pressure than healthy people.

**Figure 6:**
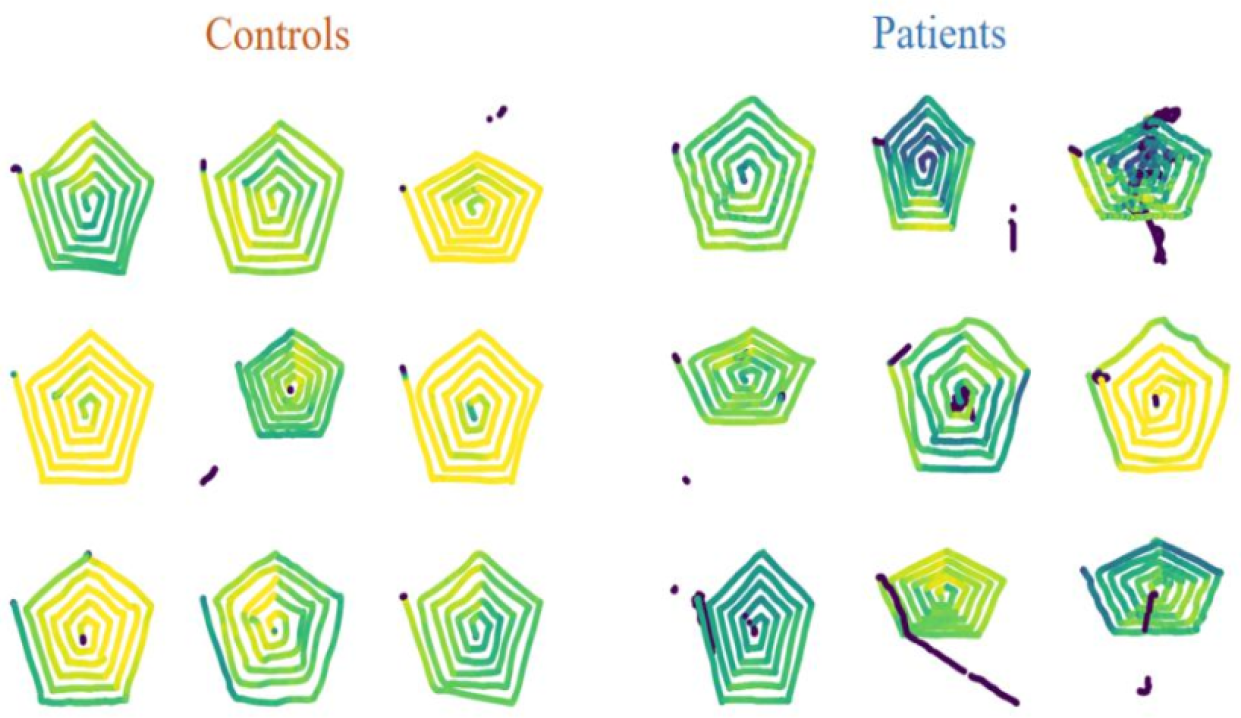
The pressure representation images of the pentagon dataset for both patient and control groups using a Viridis colour map.

#### 3.2.4 Data Representation - Time

It has been mentioned before that the speed of writing can be a characteristic that distinguishes early PD patients from healthy people. In general, PD patients usually take longer to draw for the same drawing task. Based on this feature (Scarpina et al., 2019), it is expected that including time information in the images can benefit the performance of a CNN model to distinguish PD patients from normal individuals. Fig. 7 shows the time representation images of the pentagon dataset for both the patient and control groups. A preliminary visual inspection showed us that it is difficult to distinguish the difference in the colour of the two groups with the naked eye.

**Figure 7:**
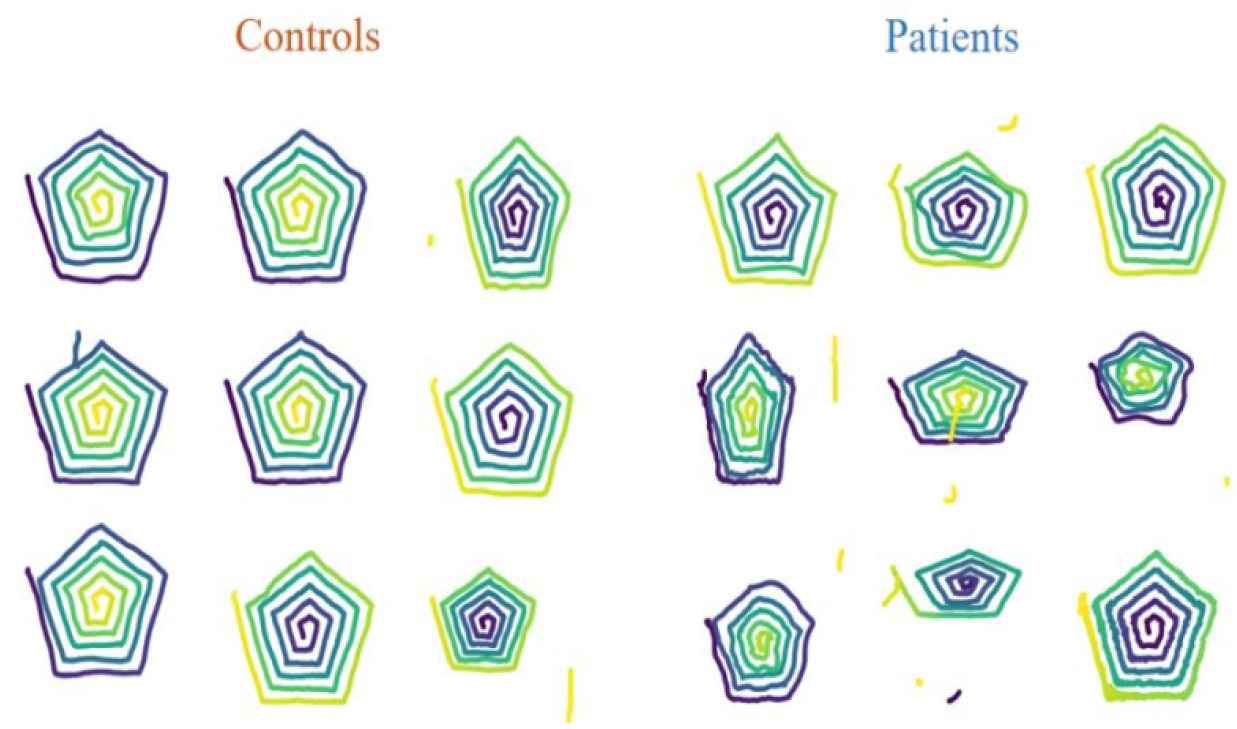
Example images from the pentagon drawing dataset using a time-based representation, where pixel intensity reflects the temporal progression of the drawing. The Viridis colour map is used to enhance visual contrast for both patient and control groups.

Although the time representation images cannot clearly show the difference between the control and the patient group, Fig. 8 helps to better understand the distribution of testing durations for the two groups. Each subject’s testing time was calculated by summing the time-steps recorded at a sampling rate of 200 Hz, where each time-step corresponds to 0.005 seconds. Half of the patients took more than 60,000 time steps (equivalent to 300 seconds), while the maximum time among the control group was only 64,003 time steps (320.015 seconds). Additionally, the mean time taken in the patient group was 49,226 time-steps (246.13 seconds) compared to 35,600 time-steps (178 seconds) in the control group, which was 13,626 time-steps (68.13 seconds) lower than the patient group.

**Figure 8:**
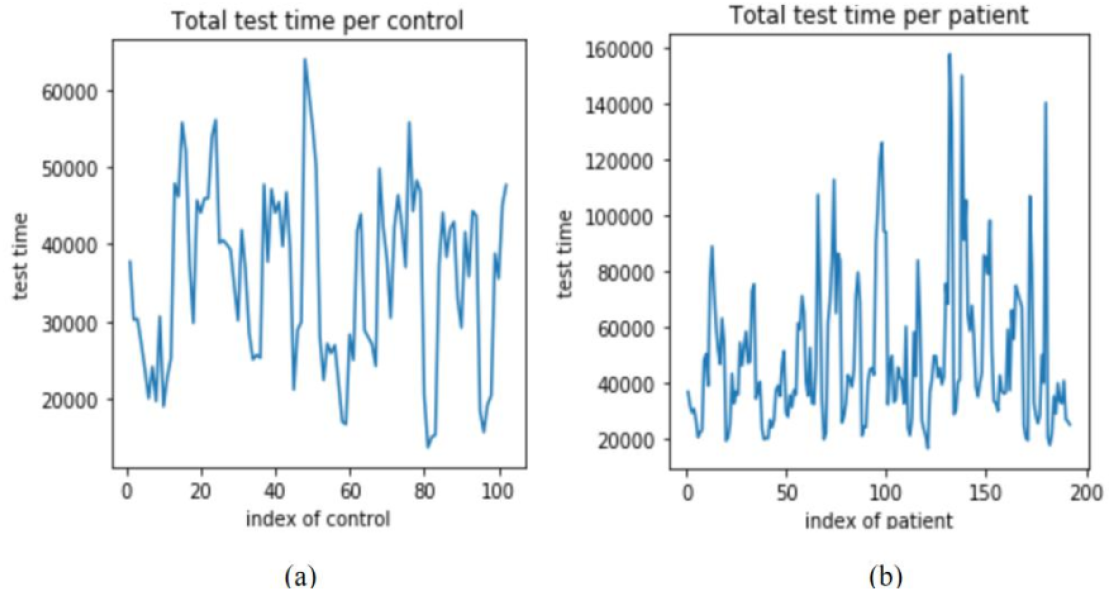
Distribution of time-steps (Y-axis) recorded for each subject (X-axis) during the drawing tasks, where (a) is the control group and (b) are the PD patients. The Y-axis represents the number of time steps captured by the digitising tablet at a constant sampling rate of 200 Hz. Each time-step corresponds to 0.005 seconds, and 20,000 time-steps represent 100 seconds of drawing activity.

In general, the above statistics also indicate that a patient’s average testing time is significantly longer than the control, which is in accordance with the study mentioned earlier. As for the time representation, images cannot show a clear difference between the two groups, which may be due to the problem of the colour mapping step, which may be an area that can be enhanced and improved afterwards.

#### 3.2.5 Data Representation - Angle

Although no study shows a correlation between the angles of the pen relative to the X and Y planes and PD, we are interested in introducing and assessing the angle representation in the image as a possible biomarker.

In the extracted signals, the angles of the pen relative to the X and Y planes are combined into a single value. The angle between the pen and its projection on the x-axis is called *θ*, which is used as the angle representation and is expressed by two variables *α* and *β* based on their spatial relationship (Fig. 9).

**Figure 9:**
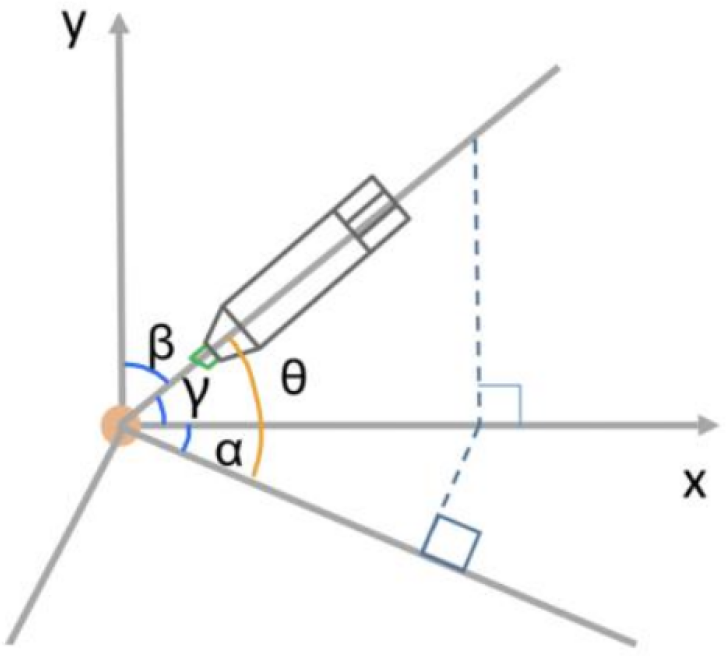
The spatial relationship among the angles of the pen.

Eq. 1 is obtained by a trigonometric calculation of *α* and *β*, and each calculated cosine value of *θ* is considered as the angle representation:

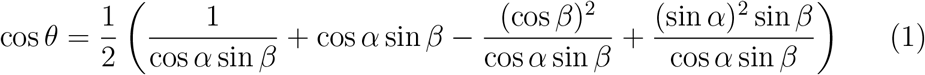

Fig. 10 shows the angle representation images of the pentagon dataset for both the patient and control groups.

**Figure 10:**
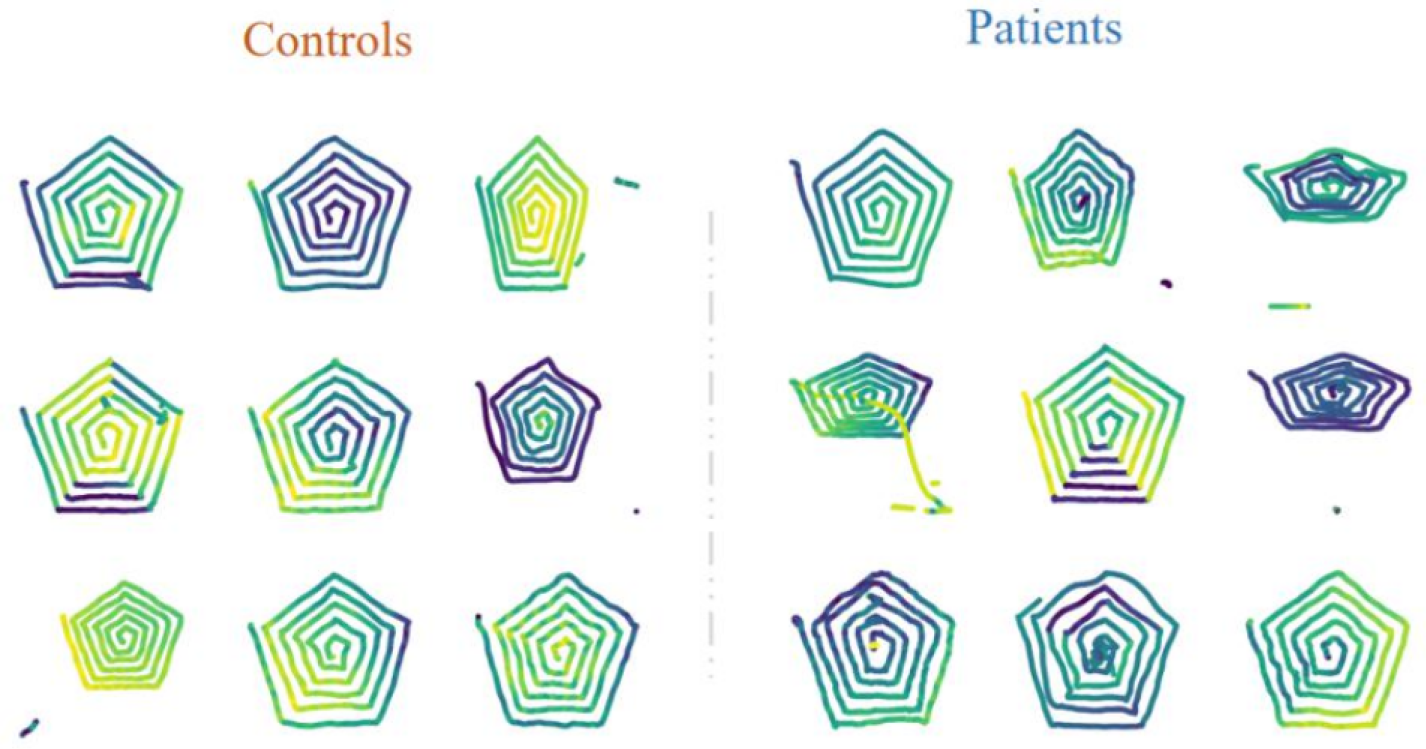
Image representations based on the pen’s orientation (tilt) captured during pentagon drawing tasks, using angle data (X, Y, Z axes) from the Wacom tablet. Examples are shown for both patient and control groups.

As for angle representation images, it is also difficult to distinguish between the two groups of drawings through simple visual inspection alone, without the use of analytical tools. However, a CNN can be expected to learn the underlying features to distinguish controls from patients. After the data representation step, the images are resized to a 32 *×* 32 resolution.

### 3.3 CNN Architecture and Evaluation Parameters

The CNN architecture used in this work was proposed by Alissa et al. (2022). It consists of two convolutional layers with 32 filters, followed by two convolutional layers with 64 filters and another two convolutional layers with 128 filters, three max-pooling layers of size (2*×* 2), six dropout layers, three dense layers and one flattened layer. All activation functions are ReLU (Rectified Linear Unit), except for the last dense layer, where a sigmoid activation function was selected to map the binary output. ReLU is the most used activation function for CNN. In each convolutional layer, we used the same padding mechanism to maintain the size of the layers after applying a series of convolutional operations. Finally, we use a stride of size (3 *×* 3). The CNN architecture is shown in Fig. 11.

**Figure 11:**
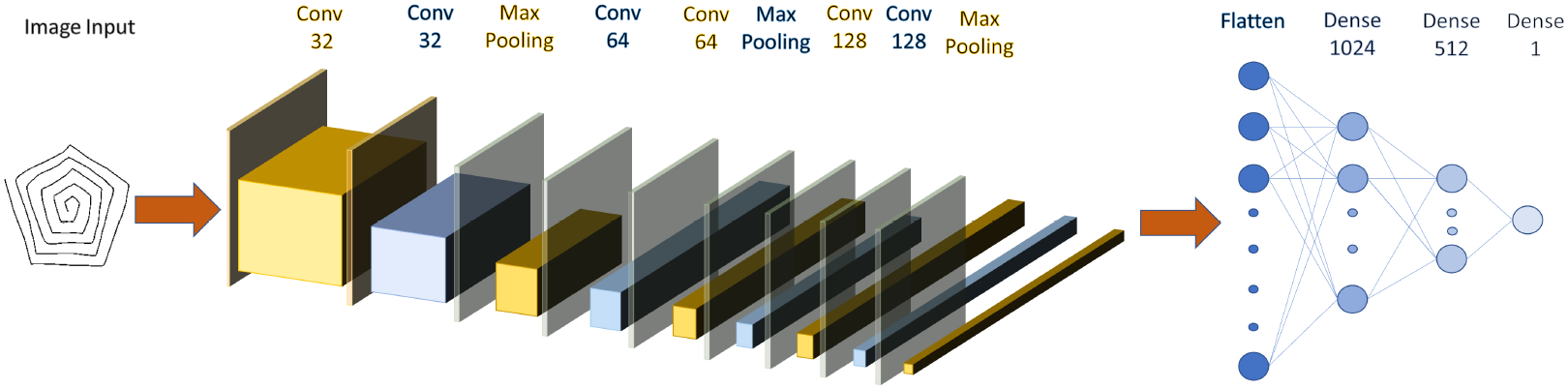
The CNN architecture used in this work (Alissa et al., 2022)

The input of the CNN model is the augmented training sets with labels. After training, the model was tested as a classifier using a test set of previously unseen images to distinguish between controls and patients. The final performance of the trained CNN model was the mean of the 10-fold cross-validation results.

The performance of the model will be evaluated using metrics such as accuracy, specificity, sensitivity, and Kappa. Except for Kappa, all are based on predicted and actual values that the system classifies as true positive (TP), true negative (TN), false positive (FP) and false negative (FN). In our work, “positive” represents patients with a label of 1, while “negative” represents controls with a label of 0. Accuracy is one of the most commonly used metrics for evaluating classification problems, which can be calculated using Eq. 2:

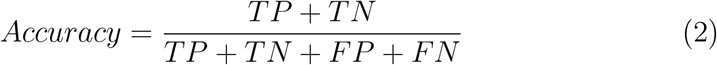

However, the accuracy measure for classification performance can be misleading, especially when using an imbalanced dataset. In contrast, Cohen’s Kappa coefficient measures agreement, correcting for the occurrence by chance, and can be applied when dealing with imbalanced datasets (Folorunso and Adeyemo, 2013). It can be calculated by Eq. 3, (Cohen, 1960):

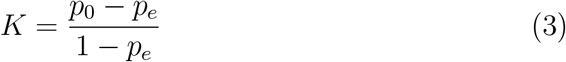

Kappa values are in the range between [− 1, 1]. There is no standard method for interpreting Kappa values, but Fleiss et al. (2013) considered that a Kappa value *>* 0.75 is excellent, between 0.4 and 0.75 is fair to good, and *<* 0.4 is a poor agreement. A Kappa value could be negative, but it is unlikely in practice.

In addition, specificity and sensitivity are also used to measure the performance of the classification results. In a diagnostic test, sensitivity measures how well a test can identify true positives, and specificity measures the true negatives. They are calculated as follows:

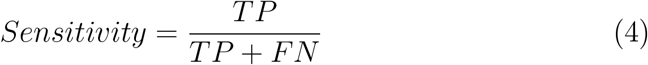

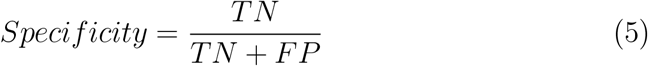

### 3.4 Data Augmentation

Due to the limited number of samples and the class imbalance in the dataset, the performance of deep learning methods can be restricted, as models may struggle to generalise well across minority classes (Johnson and Khoshgoftaar, 2019). To compensate for this limitation, an alternative is to use data augmentation techniques to artificially expand the size of the training set in both classes but oversampling the class with fewer samples, in this case, the control group, to reach the same number of instances and avoid performance deterioration associated with class imbalance (Mazurowski et al., 2008). Fig. 12 represents the flowchart that illustrates the core steps of our general experiments.

**Figure 12:**
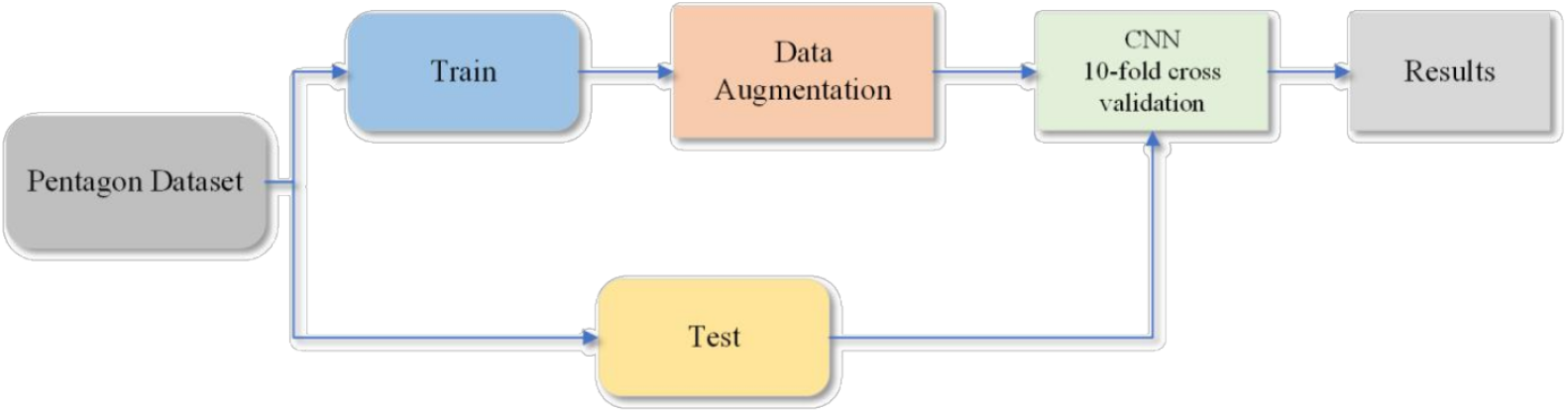
Core steps of the experiments

For data augmentation, the dataset must first be divided into the training and testing sets and then augmented with the training set. This order is essential as it ensures that the training and testing sets are statistically independent. Otherwise, results would be overoptimistic.

#### 3.4.1 Geometrical Transformations

Keras was used to replace the original batch with the new, randomly geometric transformed images using random zooming, rotation, and flipping. There are two methodologies for implementing dataset generation: one is conducted offline, whilst the other occurs online during the training of the CNN model. In this study, several factors influenced the decision to opt for the offline approach to generate new images. Primarily, the necessity to augment the sample sizes of two classes with differing multiples to balance the dataset is achievable only through offline augmentation. Furthermore, offline data augmentation facilitates displaying and storing generated images throughout the process. Consequently, the offline data augmentation method was selected for implementation. The specific steps involved in the offline data augmentation process are outlined below:

1. Load the original input image from local files.
2. Determine the strategy for geometric transformation.
3. Randomly transform the original image using a series of random translations in each batch size.
4. Secure the transformed image and save it to a designated path.
5. Repeat the two steps as mentioned earlier N times. This procedure was executed twice for the control group, whereas it occurred only once for the patient group.

It is well known that the strategy of geometric transformation plays a pivotal role in data augmentation. Techniques such as rotation, zooming, and shifting allow original images to undergo a range of modifications to enhance the dataset. However, these operations must be conducted within a reasonable range to ensure that the natural distribution of the data remains unaltered post-augmentation.

For instance, in the datasets under consideration, comprising pentagon images, most images maintain a horizontal orientation, with only a minor fraction tilting within a 5-degree range. To align the augmented data distribution closely with the original, the rotation range parameter in our geometric transformation strategy was accordingly set to 5 degrees. This careful parameterisation ensures that the augmented images reflect the inherent characteristics of the datasets, thereby maintaining the original data distribution’s integrity.

Table 2 details the specifics of the chosen strategy and outlines the parameters utilised in our geometric transformations. These parameters were meticulously selected to facilitate a balanced augmentation process, ensuring both the enhancement of the dataset and the preservation of its original distribution characteristics.

**Table 2:** Comprehensive parameters for geometrical transformations of images using Keras.

| Transformation | Parameter | Value | Description |
| --- | --- | --- | --- |
| Rotation | Rotation range | 5 | Degree range for random rotations, $[0, 5]$ |
| Zoom | Zoom range | 0.2 | Scale range for random zoom, $[0.8, 1.2]$ |
| Shift | Width shift range | 1 | Pixel range for horizontal shift, $[-1, 1]$ |
| Shift | Height shift range | 1 | Pixel range for vertical shift, $[-1, 1]$ |
| Fill | Fill mode | 'nearest' | Boundary points filled with nearest mode |

#### 3.4.2 Experiment Setup

This section delves into the experimental framework, focusing on the application of traditional geometric augmentation techniques. The experiments were meticulously designed to evaluate the performance across three distinct data representations. Initially, an imbalanced dataset for each representation was fed into the CNN to establish a baseline. Subsequently, four augmented datasets were created and separately introduced into the CNN, yielding four distinct sets of outcomes. As underscored previously, it is paramount for the training and testing datasets to be statistically independent from the outset. This independence was achieved through a randomised manual segmentation of the data. However, this method introduces a potential variability, as different segmentation approaches can significantly affect the results, a phenomenon observed in our experiments.

To mitigate the impact of segmentation variability on the experimental outcomes, we adopted a 4-fold cross-validation strategy. Illustrated in Fig. 13, this approach entails dividing the entire dataset into four distinct and mutually exclusive segments, or “folds.” In each iteration of the experiment, one fold is designated as the test set. In contrast, the remaining three folds are amalgamated and utilised as the training set, then subjected to data augmentation. This procedure ensures that each fold serves as the test set exactly once, allowing for a comprehensive evaluation of the model’s performance across the entire dataset. Within each training phase, a 10-fold cross-validation technique is employed further to refine our understanding of the model’s capabilities. This nested cross-validation strategy facilitates a more robust and nuanced analysis, culminating in an aggregated metric that reflects the mean of the model’s skill scores. By implementing a 4-fold experiment, we effectively minimise the variance in outcomes attributable to different training-test splits, thereby enhancing the reliability and interpretability of our results.

**Figure 13:**
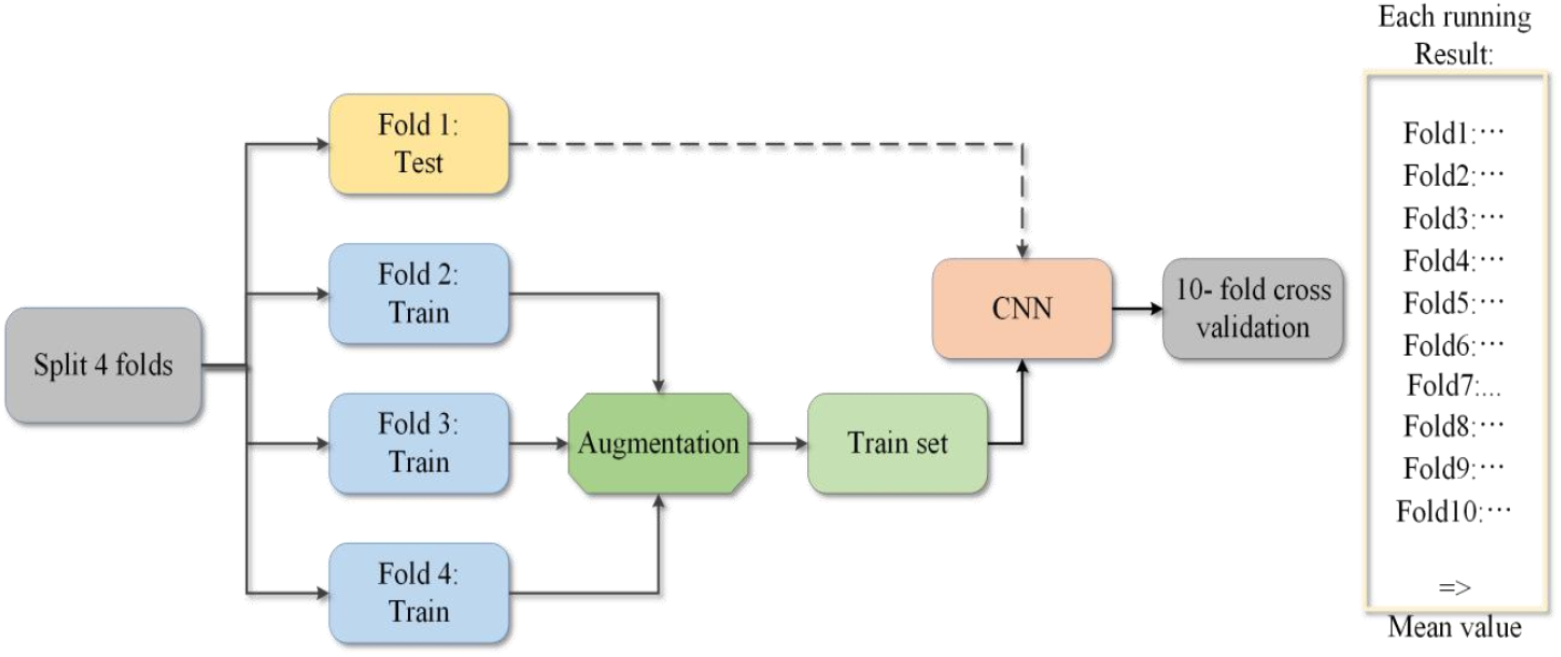
The process of the 4-fold experiment.

#### 3.4.3 Generative Adversarial Networks-based Data Augmentation

GANs represent a class of generative models capable of producing new, synthetic content. This segment elucidates the mechanism through which GANs facilitate the generation of new images within the scope of our current study, as illustrated in Fig. 14. At the heart of a GAN are two distinct but interrelated sub-models: the generator and the discriminator.

**Figure 14:**
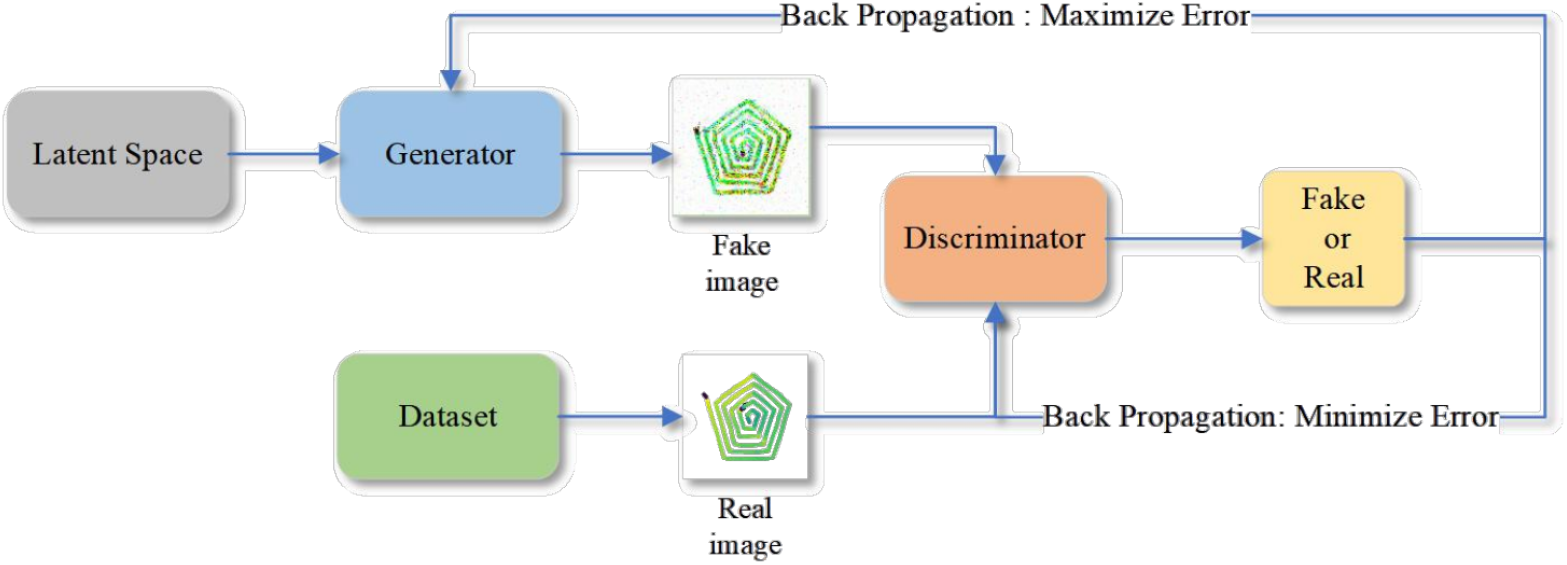
Representation of the GAN architecture used in this paper.

**Figure 15:**
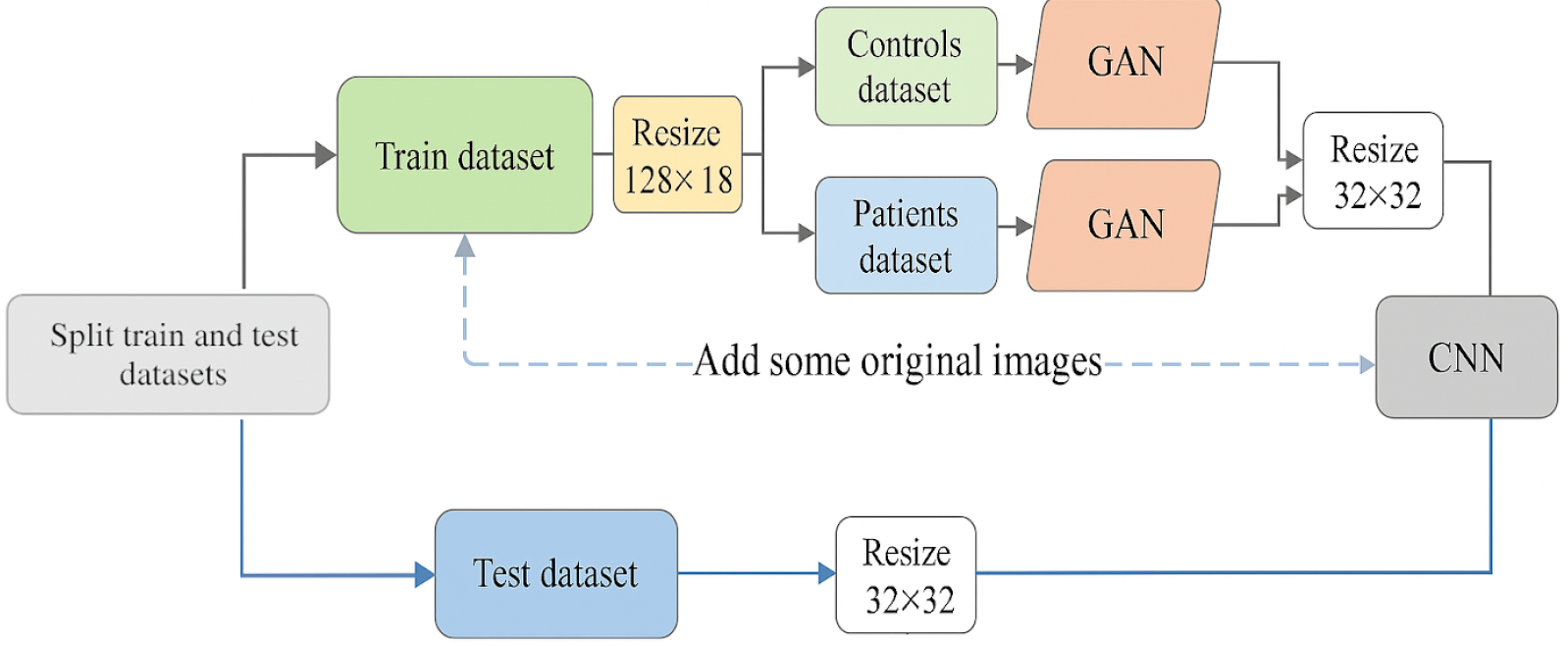
Full machine learning pipeline.

The generator’s primary objective is synthesising new data instances indistinguishable from actual, authentic data. Essentially, it strives to deceive the discriminator by producing increasingly realistic examples. Conversely, the discriminator’s role is to discern between genuine data sourced from the original domain and counterfeit data fabricated by the generator. Through this dynamic, the discriminator endeavours to minimise the classification error by accurately distinguishing real from fake.

The interaction between these two models is akin to a strategic contest, where each model continually refines its strategies to outperform the other. From the game theory perspective, this interaction seeks an equilibrium state wherein the generator produces data that perfectly mirrors the targeted distribution, making it challenging for the discriminator to make accurate judgments. At this equilibrium, the discriminator’s ability to classify data as *real* or *generated* should approximate a probability of 5 for any given input, signifying that it can no longer reliably differentiate between the two (Wang et al., 2017).

This competitive yet cooperative relationship drives both networks towards their respective goals, facilitating a process of mutual improvement. This unique dynamic underpins the efficacy of GANs in generating high-quality, diverse datasets for enhanced machine learning applications.

#### 3.4.4 Generative Adversarial Networks-based Machine Learning Pipeline

The training and testing sets must be statistically independent from the outset to ensure robust and unbiased model training. This independence is achieved by manually and randomly dividing the dataset into separate training and testing sets. After this initial division, the training set images are resized to 128 *×* 128 pixels to capture more detailed information, which results in clearer generated images. Conversely, the test set images are resized to 32*×* 32 pixels to maintain evaluation consistency.

Different experimental groups, such as controls and patients, are labelled distinctly and processed separately in the GAN to generate images. Once the GAN is trained, the generated images from both groups are combined into a single dataset. To ensure parity between the training and testing sets, the training images are resized to match the 32 pixel dimension of the test set. The training set is then expanded and balanced using the GAN before being fed into the CNN model we developed (Fig. 16).

**Figure 16:**
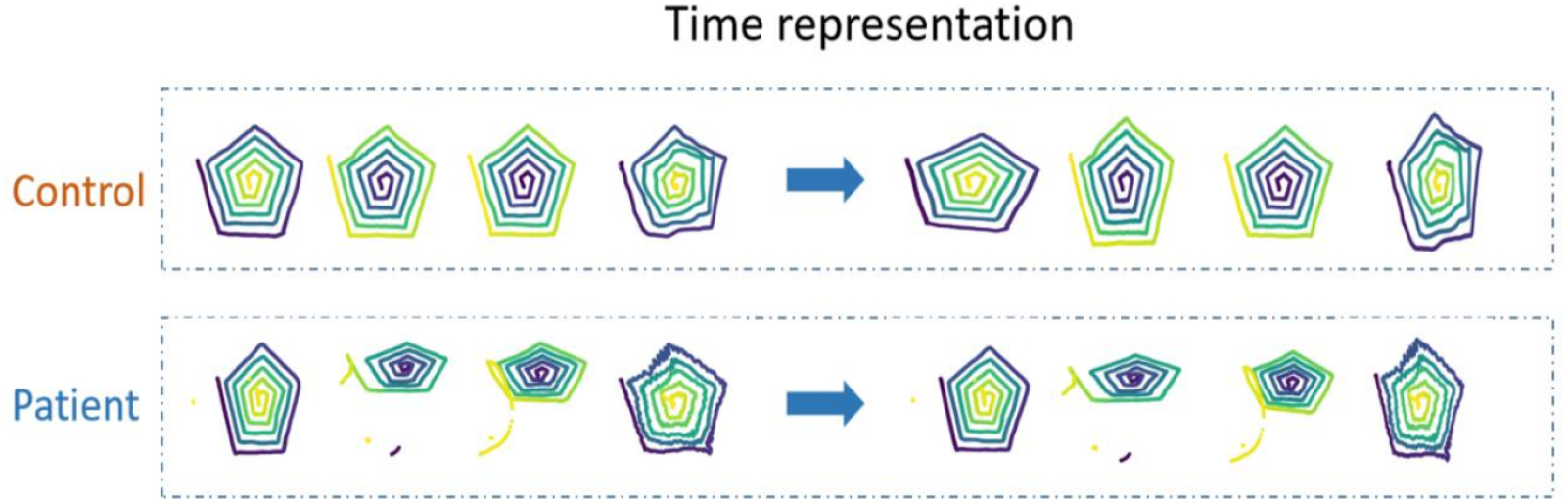
The original time representation images (right) and the augmented samples (left) were created by applying traditional geometric transformation methods.

The GAN’s learning process involves capturing more features and patterns over multiple epochs. This process is complex and time-consuming rather than linear. To facilitate the evaluation of the GAN’s progress, parameters such as the running cutoff epoch and sampling interval are set to update the generated images at specific epochs, allowing for visual assessment of the learning effect at certain points.

This entire pipeline is applied to three representations: time, pressure, and angle. Consequently, the results of the experiment are compared in two dimensions: the effectiveness of the GAN-based data augmentation across the three representations and the comparison between different augmentation methods.

### 3.5 DCGAN-based Data Augmentation method

GANs use relatively simple, fully connected networks for both the generator and discriminator models. In contrast, DCGANs employ convolutional layers in the discriminator and transposed convolutional layers in the generator. Since convolutional networks are more suitable for learning the features of images, DCGANs are better suited for this task than traditional GANs. Our subsequent work also confirms this point.

Additionally, the architectural choices in DCGANs contribute to more stable training processes, reducing the likelihood of mode collapse and leading to more reliable results. Convolutional layers enable better feature extraction and hierarchical learning, capturing complex structures in images that fully connected layers may miss (Radford et al., 2015). This ability allows DCGANs to learn more meaningful and interpretable representations of the data, which is beneficial for various downstream tasks (Odena et al., 2017).

Although GANs’ simpler network architecture results in lower learning costs compared to DCGANs, as training progresses to larger epochs, the noise in the generated images increases significantly. This increased noise can negatively impact the performance of CNNs using these images. On the other hand, images generated by the DCGAN-based data augmentation method exhibit significantly less noise, thereby enhancing the performance and reliability of the CNN results.

#### 3.5.1 Detailed Procedure of DCGAN Experiments

The procedure for the DCGAN experiments closely mirrors that of the GAN experiments, with two key differences. Firstly, for the DCGAN experiments, the training set images are resized to 64 *×* 64 pixels instead of 128 *×* 128 pixels. This adjustment reduces the computational complexity and accelerates the runtime, using the efficiency of DCGANs while still capturing essential image features.

Secondly, the original images are fed into different networks—either the traditional GAN or the DCGAN—to generate new images. This distinction ensures that the generated images reflect the unique characteristics and capabilities of each network type. Despite these differences, all other processes remain identical, allowing for a more direct and effective comparison of the results from the two experiments.

To evaluate the performance across different representations, the experiments are conducted over various epochs using the DCGAN-based data augmentation method. This approach enables testing the results of different representations, such as time, pressure, and angle, providing a comprehensive analysis of the DCGAN’s effectiveness. Maintaining consistency in all other aspects of the experimental procedure allows us to accurately assess the impact of the network architecture on the quality and utility of the generated images.

### 4. Results

#### 4.1 Results of the Original Dataset Using Different Colourmaps

This section presents the CNN performance of the original dataset across three representations using different colourmaps. As discussed in Section 3.2.2, colourful images are expected to enhance performance compared to greyscale images. A detailed comparison of this factor is provided in Table 4.

Table 3 demonstrates that the performance of the Viridis colour map slightly outperforms the Greys colour map for all three representations. Specifically, the accuracy of the classifier trained with the Viridis colour map is 3-5% higher than that trained with the Greys colour map, except for the angle representation. Additionally, the Kappa value for the pressure representation with the Viridis colour map shows a significant increase of 0.12 compared to the Greys colour map.

**Table 3:**
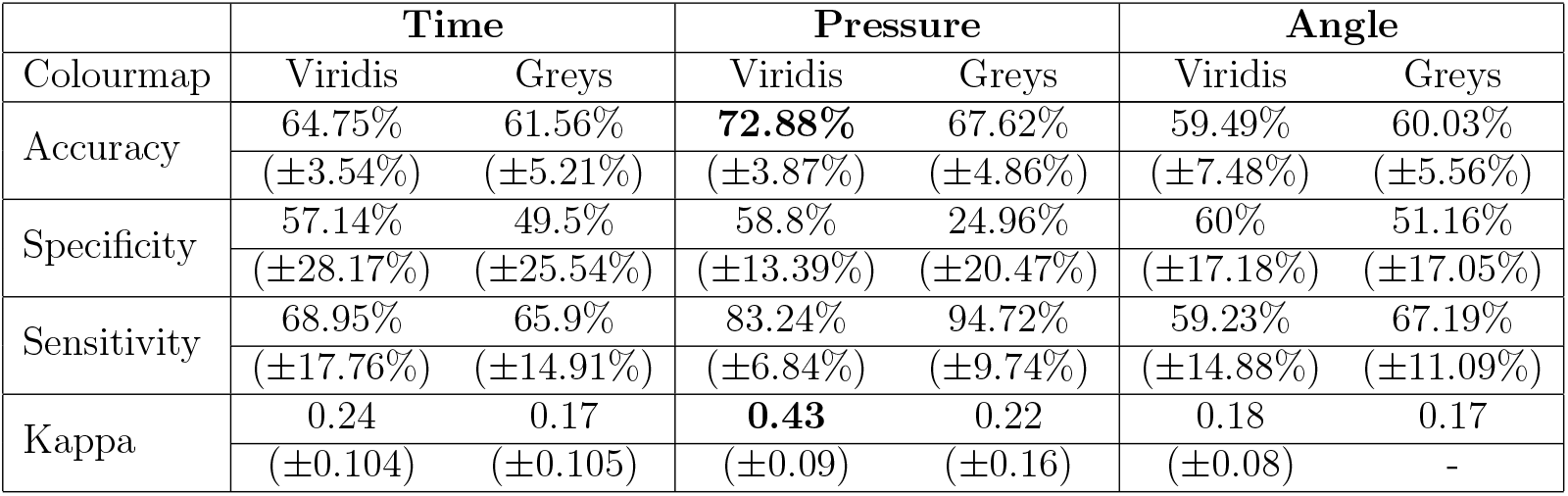
Comparison of two colour maps without data augmentation.

**Table 4:** CNN results of time representation.

| Time Representation |  |  |  |  |  |  |
| --- | --- | --- | --- | --- | --- | --- |
| Experiments | No_Aug | Fold 1 | Fold 2 | Fold 3 | Fold 4 | CV |
| Training Set | 82,154 | 304,284 | 301,310 | 300,309 | 302,311 | - |
| Test Set | 20,38 | 25,48 | 26,48 | 26,48 | 26,48 | - |
| Accuracy | 64.75% | 69.32% | 67.43% | 64.46% | 63.15% | 66.01% |
| Mean (SD) | ($\pm 3.54\%$) | ($\pm 5.13\%$) | ($\pm 5\%$) | ($\pm 4.06\%$) | ($\pm 4.92\%$) | ($\pm 4.78\%$) |
| Specificity | 57.14% | 76.4% | 60.38% | 55% | 46.4% | 59.54% |
| Mean (SD) | ($\pm 28.17\%$) | ($\pm 7.68\%$) | ($\pm 20.36\%$) | ($\pm 14.8\%$) | ($\pm 15.51\%$) | ($\pm 14.59\%$) |
| Sensitivity | 68.95% | 65.62% | 71.25% | 69.58% | 71.88% | 69.59% |
| Mean (SD) | ($\pm 17.76\%$) | ($\pm 10\%$) | ($\pm 13.1\%$) | ($\pm 11.57\%$) | ($\pm 14.14\%$) | ($\pm 12.2\%$) |
| Kappa | 0.24 | 0.38 | 0.3 | 0.24 | 0.18 | 0.27 |
| Mean (SD) | ($\pm 0.104$) | ($\pm 0.082$) | ($\pm 0.106$) | ($\pm 0.073$) | ($\pm 0.571$) | ($\pm 0.21$) |

These results indicate that the Viridis colour map is more suitable for the pressure and time representations, as it enhances the performance metrics considerably. However, for the angle representation, different colour maps do not significantly affect performance. Based on these findings, subsequent experiments were conducted using the Viridis colour map to use its benefits in improving classifier accuracy and reliability.

#### 4.2 Time Representation Results

Fig. 16 illustrates the original and generated images of the pentagon dataset in terms of time representation. This visualisation uses a colour map to represent time in figures generated from the time series data collected using the tablet. Four images are presented for comparison before and after augmentation for both control and patient groups.

As seen in the same figure, the images generated through traditional geometric transformations exhibit random zooming in and out as well as tilting. These transformations can be randomly combined during the implementation of our data augmentation strategy, enhancing the variability and robustness of the dataset.

According to the experimental process described before, five experiments were conducted using the time representation. The first experiment, serving as the control, utilised only the original time representation dataset without any augmentation, and its results are presented in the first column of Table 4. The subsequent four columns display the results of four additional experiments, each conducted using a 4-fold cross-validation approach.

The results for the time representation indicate a notable improvement in the CNN model’s performance with data augmentation and cross-validation. The accuracy of the model without augmentation is 64.75%, while the cross-validation results show a slightly higher average accuracy of 66.01%. Although the improvement is modest at 1.26%, the reduced standard deviation in cross-validation results (4.78%) compared to the non-augmented experiment (3.54%) suggests more consistency across different data folds. Specificity, measuring the true negative rate, also shows more stability, where the standard deviation drops significantly from 28.17% (No Aug) to 14.59%. This indicates a more reliable detection of true negatives. Sensitivity, the true positive rate, improves slightly from 68.95% (No Aug) to 69.59% cross-validation with a reduced standard deviation, highlighting more consistent performance. The Kappa statistic, reflecting the agreement between predicted and actual classifications, improves marginally from 0.24 (No Aug) to 0.27, with a lower standard deviation, indicating a more reliable classification performance with augmented data.

### 4.3 Pressure Results

Similarly, Fig. 17 shows the original images and the generated images of the pentagon dataset in terms of pressure representation, where pressure refers to the force exerted by the subject when drawing with a pen on the tablet.

**Figure 17:**
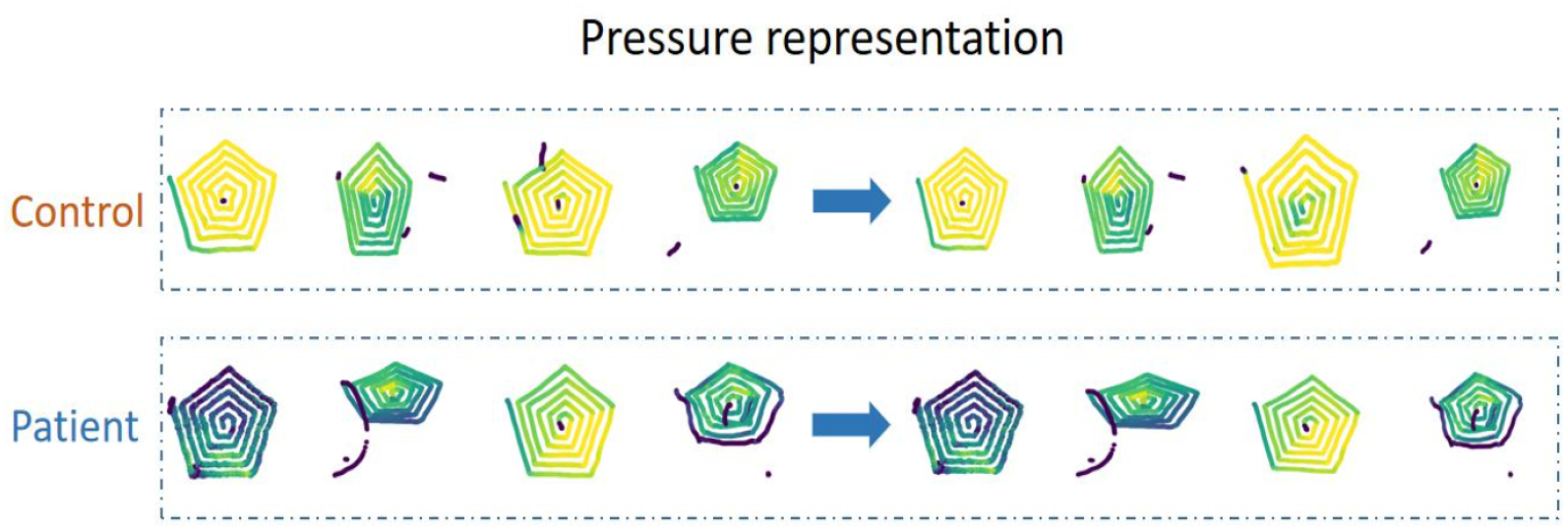
The original images (right) and the images augmented by traditional geometric transformation methods (left) in terms of pressure representation. Transformations include random zooming, rotation, and flipping.

Table 5 presents the CNN results for pressure representations. The table compares the performance metrics of models trained on the original dataset without augmentation, models trained on an augmented dataset using geometric transformations, and 4-fold cross-validation.

**Table 5:** CNN results of pressure representation.

| Pressure Representation |  |  |  |  |  |  |
| --- | --- | --- | --- | --- | --- | --- |
| Experiments | No_Aug | Fold 1 | Fold 2 | Fold 3 | Fold 4 | CV |
| Training Set | 82,154 | 307,310 | 303,310 | 302,309 | 307,310 | - |
| Test Set | 2,038 | 2,548 | 2,548 | 2,648 | 2,648 | - |
| Accuracy | 72.88% | 80.14% | 72.7% | 75.41% | 70.82% | 74.77% |
| Mean (SD) | ($\pm 3.87\%$) | ($\pm 3.53\%$) | ($\pm 3.19\%$) | ($\pm 3.57\%$) | ($\pm 2.74\%$) | ($\pm 3.26\%$) |
| Specificity | 58.8% | 78.4% | 59.23% | 56.15% | 58.8% | 59.54% |
| Mean (SD) | ($\pm 13.39\%$) | ($\pm 8.43\%$) | ($\pm 9.76\%$) | ($\pm 10.06\%$) | ($\pm 12.53\%$) | ($\pm 10.19\%$) |
| Sensitivity | 83.24% | 81.04% | 80% | 85.83% | 77.08% | 80.99% |
| Mean (SD) | ($\pm 6.84\%$) | ($\pm 6.07\%$) | ($\pm 3.86\%$) | ($\pm 5.42\%$) | ($\pm 5.82\%$) | ($\pm 5.29\%$) |
| Kappa | 0.43 | 0.57 | 0.39 | 0.44 | 0.35 | 0.27 |
| Mean (SD) | ($\pm 0.104$) | ($\pm 0.082$) | ($\pm 0.106$) | ($\pm 0.073$) | ($\pm 0.571$) | ($\pm 0.44$) |

In the case of pressure representation, the model without augmentation shows an accuracy of 72.88%, which improves to 74.77%, accompanied by a standard deviation of 3.26%. This reflects a moderate enhancement in classification accuracy with data augmentation. The specificity shows a minor improvement in mean values, from 58.8% (No Aug) to 59.54%, but the notable reduction in standard deviation from 13.39% to 10.19% suggests more consistent performance in identifying true negatives. Sensitivity remains high in both scenarios, with a slight decrease from 83.24% (No Aug) to 80.99%, balanced by a lower standard deviation, ensuring consistent identification of positive cases. The Kappa statistic shows a slight increase from 0.43 (No Aug) to 0.44, indicating improved reliability in the classification results.

### 4.4 Angle Results

Building on the previous section’s discussion of pressure representation, Fig. 18 illustrates the original images and the generated images of the pentagon dataset in terms of angle representation. Here, the angle refers to the combined angle of the three axes represented by the pen and the tablet.

**Figure 18:**
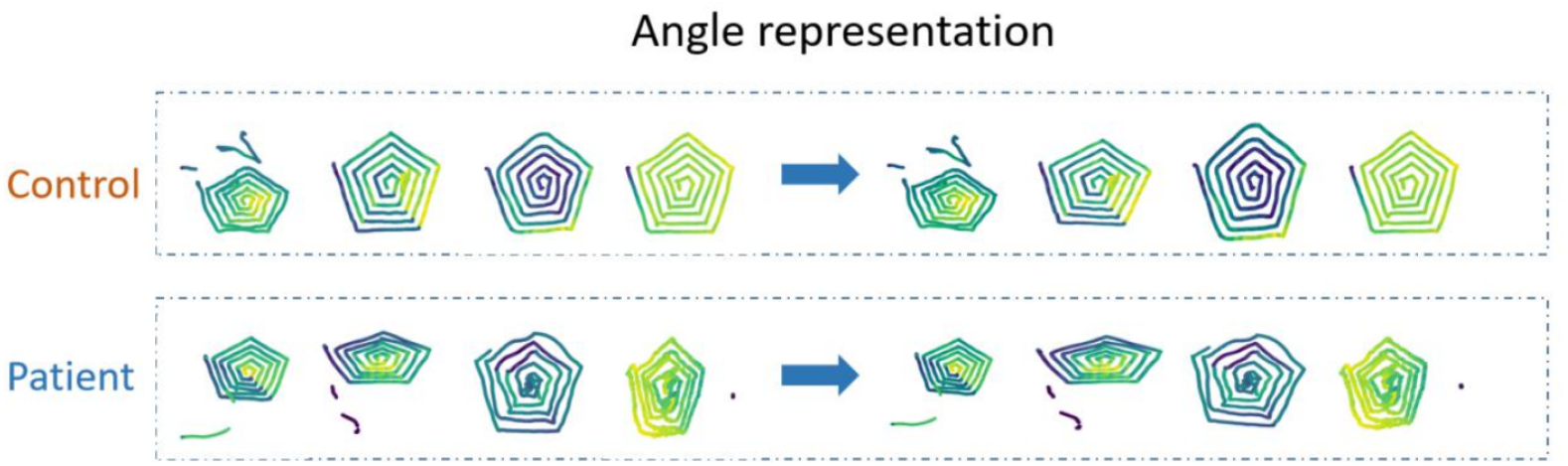
The original images (right) and the images augmented by traditional geometric transformation methods (left) in terms of angle representation.

Table 6 presents the CNN results for pressure representations. The table compares the performance metrics of models trained on the original dataset without augmentation, models trained on an augmented dataset using geometric transformations, and 4-fold cross-validation.

**Table 6:** CNN results of angle representation.

| Angle Representation |  |  |  |  |  |  |
| --- | --- | --- | --- | --- | --- | --- |
| Experiments | No_Aug | Fold 1 | Fold 2 | Fold 3 | Fold 4 | CV |
| Training Set | 82,154 | 227,224 | 226,232 | 223,237 | 226,232 | - |
| Test Set | 2,038 | 2,548 | 2,548 | 2,648 | 2,648 | - |
| Accuracy | 59.49% | 62.74% | 66.22% | 63.65% | 52.47% | 61.27% |
| Mean (SD) | ($\pm 7.48\%$) | ($\pm 4.58\%$) | ($\pm 4.05\%$) | ($\pm 3.33\%$) | ($\pm 5.94\%$) | ($\pm 4.47\%$) |
| Specificity | 60% | 49.2% | 71.15% | 47.69% | 55.2% | 55.81% |
| Mean (SD) | ($\pm 17.18\%$) | ($\pm 17.62\%$) | ($\pm 7.35\%$) | ($\pm 16.06\%$) | ($\pm 11.97\%$) | ($\pm 13.25\%$) |
| Sensitivity | 59.23% | 69.79% | 63.54% | 72.29% | 51.04% | 64.16% |
| Mean (SD) | ($\pm 14.88\%$) | ($\pm 10.99\%$) | ($\pm 6.13\%$) | ($\pm 7.63\%$) | ($\pm 5.98\%$) | ($\pm 7.68\%$) |
| Kappa | 0.18 | 0.18 | 0.32 | 0.2 | 0.055 | 0.19 |
| Mean (SD) | ($\pm 0.123$) | ($\pm 0.106$) | ($\pm 0.076$) | ($\pm 0.101$) | ($\pm 0.124$) | ($\pm 0.102$) |

For angle representation, the model accuracy improves modestly from 59.49% (No Aug) to 61.27% (with augmentation), with a standard deviation of 4.47%. The consistency introduced by the augmented data is evident in the range of accuracy (52.47% to 66.22%), highlighting robustness. Specificity shows mixed results, with the No Aug experiment having a mean specificity of 60% (SD = 17.18%) compared to 55.81% (SD = 13.25%), suggesting more consistent performance despite a slight reduction in mean values. Sensitivity improves with augmentation, with a mean value of 64.16% and a lower standard deviation of 7.68%, compared to 59.23% (SD = 14.88%) for No Aug. The Kappa statistic improves marginally from 0.18 (No Aug) to 0.19, with lower variability, emphasising the benefits of augmentation in providing reliable performance assessments.

Overall, across time, pressure, and angle representations, data augmentation and cross-validation consistently enhance model performance. These techniques lead to improvements in accuracy, sensitivity, and Kappa statistics while ensuring more stable and reliable results, as reflected by lower standard deviations. This underscores the importance of incorporating data augmentation and cross-validation in developing robust and reliable CNN models, especially in scenarios with limited original data.

### 4.5 Results of GAN-based Data Augmentation Method

The images generated by GANs for the two groups (control and patient) across various epochs are shown in Fig.19, providing an intuitive illustration of the GAN learning process. The CNN performance metrics for different representations are detailed in Table 7. For this example, images from the patient group are highlighted to illustrate the progression. In the GAN architecture, the generator model begins by taking a random latent space input, typically a 100-dimensional vector where each variable follows a Gaussian distribution with a mean of zero and a standard deviation of one (Brownlee, 2019), corresponding to the initial image at epoch 0.

**Figure 19:**
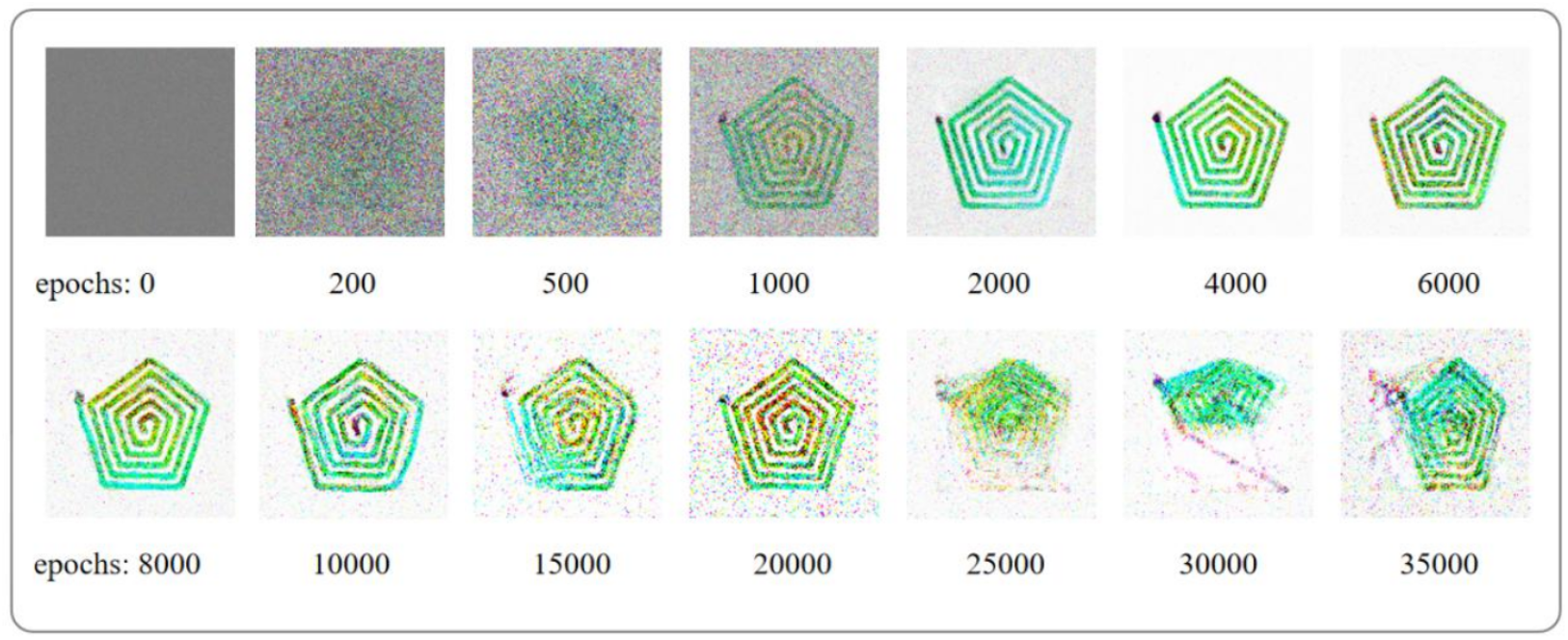
Examples of images generated by GANs for the patient group at various epochs, illustrating the progression of learned features (Pressure representation).

**Table 7:** Performance of CNN on three representations at two GAN training epochs using GAN-based data augmentation.

|  | Representations |  |  |  |  |  |
| --- | --- | --- | --- | --- | --- | --- |
|  | Time |  | Pressure |  | Angle |  |
| GAN epochs | 6,000 | 8,000 | 6,000 | 8,000 | 6,000 | 8,000 |
| Train set | 272,261 | 273,261 | 269,261 | 261,262 | 273,261 | 261,272 |
| Test set | 21,39 | 21,39 | 21,39 | 21,39 | 21,39 | 21,39 |
| Accuracy | 61.17%<br>(+/-6.99%) | 61.17%<br>(+/-5.38%) | 71.50%<br>(+/-4.74%) | 72.83%<br>(+/-5.78%) | 59.45%<br>(+/-5.66%) | 60.33%<br>(+/-6.36%) |
| Specificity | 70%<br>(+/-22.14%) | 65.71%<br>(+/-16.61%) | 70.48%<br>(+/-11.02%) | 76.19%<br>(+/-7.38%) | 61.76%<br>(+/-9.28%) | 62.86%<br>(+/-11.43%) |
| Sensitivity | 56.41%<br>(+/-20.06%) | 58.72%<br>(+/-14.48%) | 72.05%<br>(+/-9.49%) | 71.03%<br>(+/-9.53%) | 58.21%<br>(+/-10.61%) | 58.97%<br>(+/-7.43%) |
| Kappa | 0.24<br>(+/-0.09) | 0.22<br>(+/-0.08) | 0.41<br>(+/-0.083) | 0.44<br>(+/-0.101) | 0.19<br>(+/-0.091) | 0.20<br>(+/-0.128) |

As training progresses, the images generated by the GAN evolve. By epoch 1000, the generated images start to show a blurry pentagon shape and initial colour patterns, though significant noise remains around the subject. At 2000 epochs, the boundary between the subject and the background becomes clearer, and noise is noticeably reduced. By epoch 8000, the images display a broader range of colours and clearer shapes, indicating an improvement in the GAN’s ability to generate varied yet structured representations. However, as training continues past 8000 epochs, the images begin to exhibit an increase in noise.

At 25,000 epochs, the generated images appear notably different, becoming more blurred with less defined boundaries. This change suggests that the GAN’s generator is exploring additional patterns and features in the data, with the images at 30,000 and 35,000 epochs displaying 3-4 distinct patterns. This variation may reflect the generator’s attempt to capture more complex aspects of the input data, though with some loss of clarity.

For the generated images of the control group across different epochs (Fig. 20), a similar pattern is observed to that of the patient group, with consistent feature learning up to around 8000 epochs. This similarity suggests that the GAN’s generator successfully captures the control images’ core features within this range. However, between 10,000 and 30,000 epochs, the generated images display unusual colour variations that deviate significantly from the original images. Interestingly, by 35,000 epochs, the colours appear to stabilize and more closely resemble the original images. This fluctuation in generated images highlights the non-linear nature of GAN training, emphasizing the need for careful monitoring and patience throughout the learning process.

**Figure 20:**
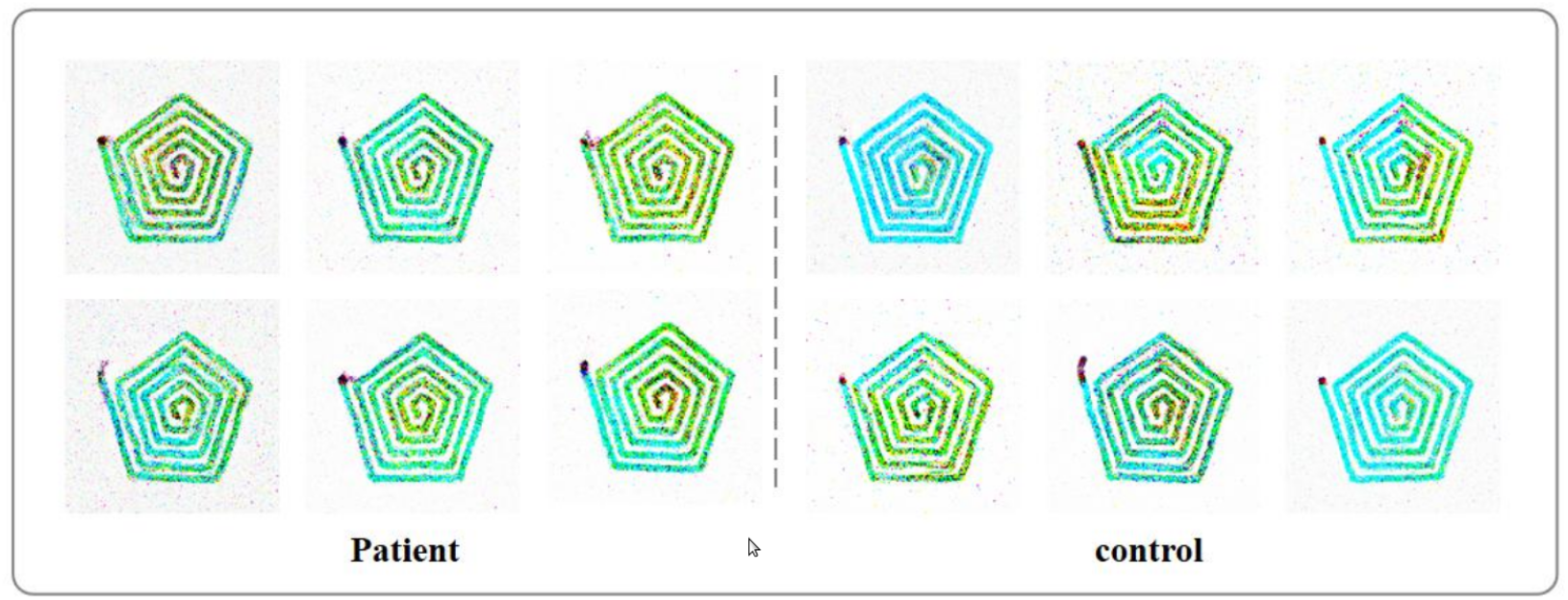
Examples of images generated by GANs for the control group at various epochs, illustrating the progression of learned features (Pressure representation).

Table 7 presents the CNN results for all three representations at 6000 and 8000 epochs using GAN-based data augmentation. It details the number of epochs, the size of the augmented training set (for both patient and control groups), the size of the test set, and four key evaluation metrics for the trained CNN model. The table shows that the pressure representation demonstrates the best performance among the three representations, with approximately 72% accuracy and a Kappa value of 0.41. In contrast, the other two representations, time and angle, achieved around 60% accuracy and a Kappa value of 0.20. Additionally, there is minimal difference in CNN performance between the GAN models trained for 4,000 and 6,000 epochs, indicating that increasing epochs beyond this range may yield diminishing returns in terms of accuracy.

As shown in Fig. 21, the images generated by GANs for patients at 35,000 epochs exhibit a wider range of patterns compared to those generated at 8000 epochs. However, these later images also contain considerable noise around the subject, which may impact the model’s performance. Since DCGANs are generally more effective at capturing detailed features in images, it is anticipated that utilizing DCGANs could improve the results compared to standard GANs.

**Figure 21:**
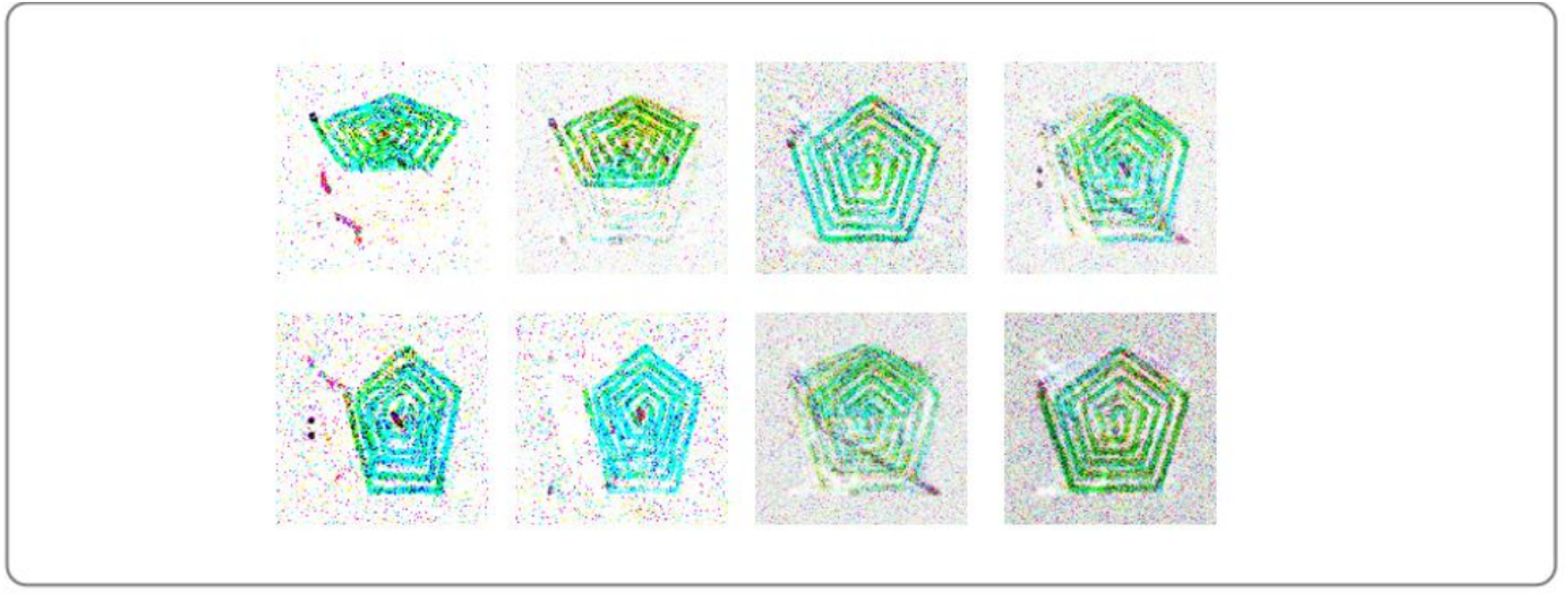
Examples of generated images for the patient group at 35,000 epochs using GANs (pressure representation), illustrating increased pattern diversity and noise.

### 4.6 Performance of DCGAN-based Data Augmentation Method

As previously mentioned, the images generated by GANs tend to accumulate noise after a certain number of training epochs. DCGANs, known for their ability to capture image features more effectively, are expected to address this issue. The generated images of the pressure representation by DCGANs for both groups at various epochs are shown in Fig. 22 and 23, providing a clear illustration of DCGANs’ learning process.

**Figure 22:**
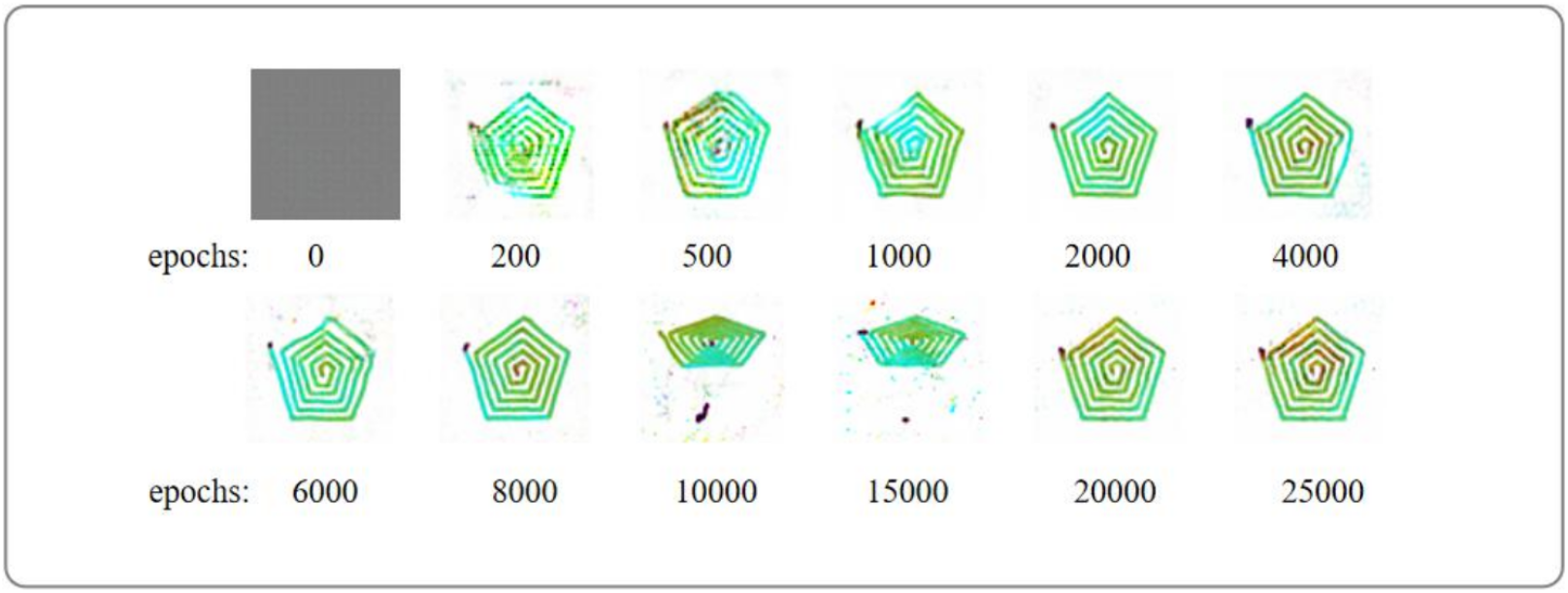
Examples of generated images for the patient group at different epochs using DCGANs (Pressure representation).

**Figure 23:**
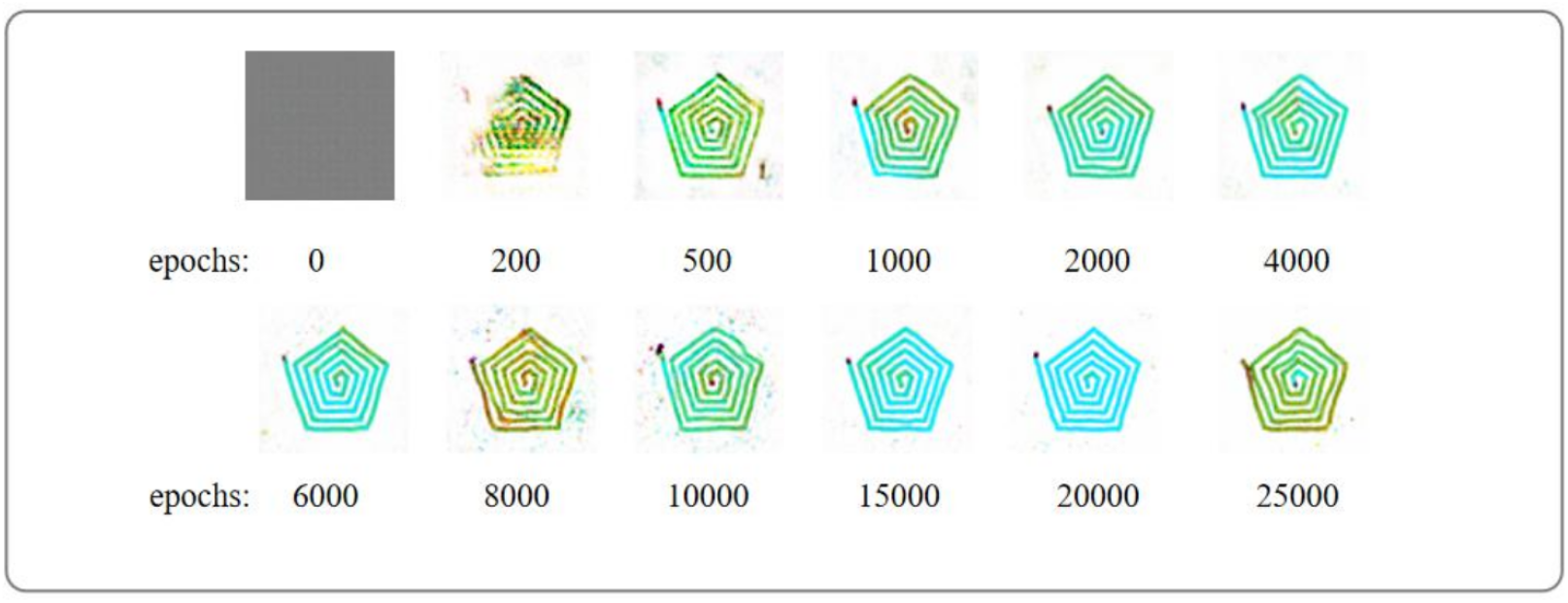
Examples of generated images for the control group at different epochs using DCGANs (Pressure representation).

In Fig. 22, the generated images for the patient group at 10,000 epochs show an early tendency to capture diverse patterns. However, the patterns in the generated images at some stages appear repetitive and could benefit from additional epochs to enhance variation. For the control group, shown in Fig. 23, generated images between 10,000 and 20,000 epochs exhibit unusual colour variations, a similar issue encountered with GANs.

Compared to GANs, the images generated by DCGANs are consistently clearer and contain less noise. Additionally, DCGANs demonstrate a faster learning speed than GANs, likely due to the more advanced network architecture. For instance, at just 500 epochs, the images generated by DCGANs already show a well-defined subject with minimal noise, whereas GANs at the same stage produce only basic shapes with significant noise. In other words, to achieve images of similar quality, DCGANs require fewer epochs than GANs. Another notable difference is observed in the shape details: while GANs tend to generate standard pentagons up to 10,000 epochs, the pentagons produced by DCGANs sometimes exhibit slight distortions along the sides, adding a degree of variability to the generated images.

Table 8 summarises the CNN results for the three representations at 4,000 and 6,000 epochs using DCGAN-based data augmentation. The table details the number of epochs, the training set size for both patient and control groups, the size of the test set, and four key evaluation metrics for the trained CNN model. Among the three representations, the pressure representation demonstrates the best performance, achieving approximately 73% accuracy and a Kappa value of 0.44. In contrast, the other two representations—time and angle—reach around 70% accuracy and a Kappa value of 0.25. Furthermore, there is minimal difference in CNN performance between the DCGAN models trained for 4,000 and 6,000 epochs, suggesting that additional epochs beyond this range may not significantly improve results.

**Table 8:** Performance of the CNN on three representations at two DCGAN training epochs using DCGAN-based data augmentation.

| Representations |  |  |  |  |  |  |
| --- | --- | --- | --- | --- | --- | --- |
|  | Time |  | Pressure |  | Angle |  |
| DCGAN epochs | 4,000 | 6,000 | 4,000 | 6,000 | 4,000 | 6,000 |
| Train set | 267,261 | 258,261 | 262,261 | 261,262 | 269,261 | 267,261 |
| Test set | 21,39 | 21,39 | 21,39 | 21,39 | 21,39 | 21,39 |
| Accuracy | 63.17%<br>(+/-5.02%) | 60.5%<br>(+/-3.08%) | 73.67%<br>(+/-4.4%) | 72.83%<br>(+/-5.78%) | 58.67%<br>(+/-4.27%) | 62.00%<br>(+/-3.93%) |
| Specificity | 67.62%<br>(+/-12.74%) | 56.19%<br>(+/-13.60%) | 71.43%<br>(+/-12.60%) | 76.19%<br>(+/-7.38%) | 75.24%<br>(+/-10.61%) | 69.52%<br>(+/-9.33%) |
| Sensitivity | 60.77%<br>(+/-8.03%) | 62.82%<br>(+/-10.83%) | 74.87%<br>(+/-8.41%) | 71.03%<br>(+/-9.53%) | 49.74%<br>(+/-10.65%) | 57.95%<br>(+/-7.79%) |
| Kappa | 0.26<br>(+/-0.106) | 0.18<br>(+/-0.042) | 0.44<br>(+/-0.088) | 0.44<br>(+/-0.101) | 0.22<br>(+/-0.054) | 0.25<br>(+/-0.068) |

### 4.7 Comparison of Results Across the Three Augmentation Methods

In this section, we extract the best-performing experiments from each augmentation method to compare their effectiveness. Table 9 presents the results of the three data augmentation methods for the time representation. The table compares four experiments, with the first column (No aug) representing the original, unaugmented time representation dataset. The original training set consists of only 82 images for controls and 154 for patients. Data augmentation expanded the training dataset to three times its original size, with balanced classes. Among these four experiments, the geometric transformation method achieved the best CNN performance, improving accuracy by 5% and increasing the Kappa value by 0.12 compared to the No aug experiment. In contrast, the performance of the DCGAN- and GAN-based augmentation methods was slightly lower than that of the No aug experiment.

**Table 9:** Comparison of results across three data augmentation methods for the time representation.

| Time Representation |  |  |  |  |
| --- | --- | --- | --- | --- |
| Augmentation Methods | No_aug | Traditional | GAN | DCGAN |
| Train set | 82,154 | 304,284 | 272,261 | 267,261 |
| Test set | 20,38 | 25,48 | 21,39 | 21,39 |
| Accuracy | 64.75%<br>(+/-3.54%) | 69.32%<br>(+/-5.13%) | 61.17%<br>(+/-6.99%) | 63.17%<br>(+/-5.02%) |
| Specificity | 57.14%<br>(+/-28.17%) | 76.40%<br>(+/-7.68%) | 70.00%<br>(+/-22.14%) | 67.62%<br>(+/-12.74%) |
| Sensitivity | 68.95%<br>(+/-17.76%) | 65.62%<br>(+/-10.00%) | 56.41%<br>(+/-20.06%) | 60.77%<br>(+/-8.03%) |
| Kappa | 0.24<br>(+/-0.104) | 0.38<br>(+/-0.082) | 0.24<br>(+/-0.090) | 0.26<br>(+/-0.106) |

Table 10 presents the results of the three data augmentation methods for the pressure representation. Similar to the previous experiment, the augmented training set is approximately three times larger than the original. The traditional data augmentation method achieves the best CNN performance among the four experiments, with a notable improvement in both accuracy and Kappa compared to the No aug experiment. Specifically, the average accuracy increases from 72.88% to 80.14%, while the Kappa value rises from 0.43 to 0.57.

**Table 10:**
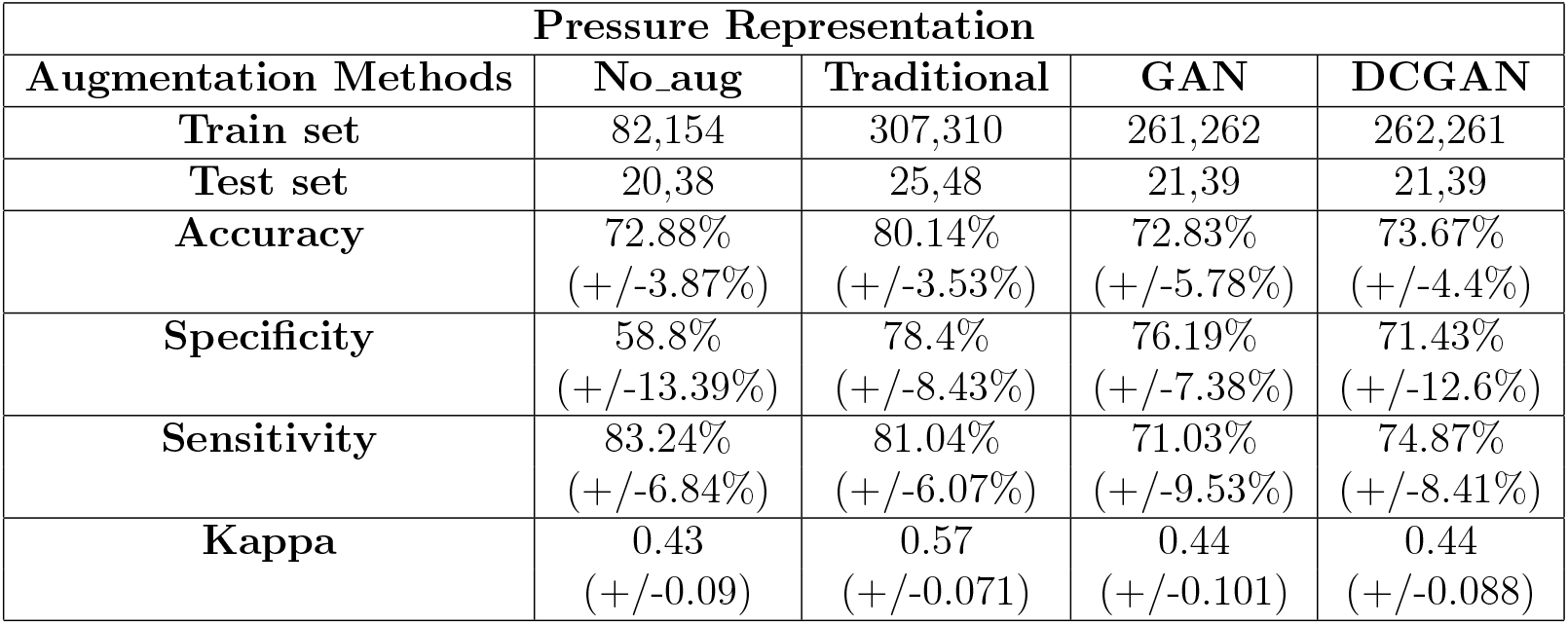
Comparison of results across three data augmentation methods for the pressure representation.

| Pressure Representation |  |  |  |  |
| --- | --- | --- | --- | --- |
| Augmentation Methods | No_aug | Traditional | GAN | DCGAN |
| Train set | 82,154 | 307,310 | 261,262 | 262,261 |
| Test set | 20,38 | 25,48 | 21,39 | 21,39 |
| Accuracy | 72.88%<br>(+/-3.87%) | 80.14%<br>(+/-3.53%) | 72.83%<br>(+/-5.78%) | 73.67%<br>(+/-4.4%) |
| Specificity | 58.8%<br>(+/-13.39%) | 78.4%<br>(+/-8.43%) | 76.19%<br>(+/-7.38%) | 71.43%<br>(+/-12.6%) |
| Sensitivity | 83.24%<br>(+/-6.84%) | 81.04%<br>(+/-6.07%) | 71.03%<br>(+/-9.53%) | 74.87%<br>(+/-8.41%) |
| Kappa | 0.43<br>(+/-0.09) | 0.57<br>(+/-0.071) | 0.44<br>(+/-0.101) | 0.44<br>(+/-0.088) |

For the GAN- and DCGAN-based augmentation methods, the accuracy and Kappa values are similar to those of the No aug experiment, showing less impact on performance compared to the traditional method. Additionally, a common outcome across these three augmented experiments is a reduction in the discrepancy between specificity and sensitivity, likely due to the more balanced dataset provided by data augmentation.

Table 11 presents the results of the three data augmentation methods for the angle representation. All three augmentation methods demonstrate significant improvements in CNN performance compared to the No aug experiment. Notably, the traditional method achieves the largest gains, with a 7% increase in accuracy and a 0.14 improvement in the Kappa value. Additionally, the DCGAN-based data augmentation method provides a moderate enhancement, with a 3% increase in accuracy and a 0.07 increase in Kappa.

**Table 11:** Comparison of results across three data augmentation methods for the angle representation.

| Angle Representation |  |  |  |  |
| --- | --- | --- | --- | --- |
| Augmentation Methods | No_aug | Traditional | GAN | DCGAN |
| Train set | 82,154 | 226,232 | 261,272 | 267,261 |
| Test set | 20,38 | 25,48 | 21,39 | 21,39 |
| Accuracy | 59.49%<br>(+/-7.48%) | 66.22%<br>(+/-4.05%) | 60.33%<br>(+/-6.36%) | 62%<br>(+/-3.93%) |
| Specificity | 60%<br>(+/-17.18%) | 71.15%<br>(+/-7.35%) | 62.86%<br>(+/-11.43%) | 69.52%<br>(+/-9.33%) |
| Sensitivity | 59.23%<br>(+/-14.88%) | 63.54%<br>(+/-6.13%) | 58.97%<br>(+/-7.43%) | 57.95%<br>(+/-7.79%) |
| Kappa | 0.18<br>(+/-0.123) | 0.32<br>(+/-0.076) | 0.20<br>(+/-0.128) | 0.25<br>(+/-0.068) |

## 5. Discussion

In this study, we explored and tested various data augmentation methods on a CNN model to automatically detect PD from figure copying tasks. These augmentation techniques expanded the limited dataset, balanced class distributions, and enhanced the model’s invariance and generalisation. Among the methods tested, traditional geometric augmentation led to the most notable improvements in performance, especially when applied to pressure-based representations of the drawing data. Nevertheless, achieving clinically meaningful performance cannot rely solely on data augmentation. A more robust solution lies in expanding the diversity of the original dataset itself, as augmented images, regardless of the method, are still derived from the same underlying patterns.

A critical component of this work was the transformation of drawing data from time-series inputs into image representations. Three novel input modalities were explored: time, pressure, and angle of the pen relative to the tablet surface. Across all augmentation strategies, the pressure representation consistently achieved the highest classification performance, followed by time and angle. This suggests that pressure signals carry more discriminative information relevant to PD-related motor impairment. Furthermore, while colour gradients were used to enhance image contrast, they also introduced increased computational complexity and unexpected artefacts in GAN-generated images, particularly within control samples. These findings raise questions about the utility of colour in synthetic augmentation, and grayscale alternatives may offer a more stable solution for generative models. Interestingly, our experiments revealed that different colourmaps had a measurable influence on CNN performance, which was unexpected and warrants further investigation.

The present study builds on and addresses methodological limitations in prior work using the same dataset. In particular, a previous CNN-based study (Alissa et al., 2022) focused on two drawing types and reported high accuracy (95%) using traditional augmentation techniques. However, that work did not account for subject-level separation in cross-validation, potentially leading to data leakage and over-optimistic performance estimates. Another study (Shenoy et al., 2021) took a more conservative approach by using a single sample per subject without augmentation and applying recurrent neural networks (LSTM and Echo State Networks), achieving accuracies of 91% and 93.7%, respectively. Our approach improves upon both by employing subject-wise cross-validation, ensuring no data leakage, and using all available samples per subject. Though our CNN results, particularly with time and angle representations, did not match the inflated accuracy from (Alissa et al., 2022), they are more realistic and robust for deployment scenarios.

The observed drop in performance under proper validation (e.g., 80.14% accuracy for pressure + traditional augmentation, compared to 95% in (Alissa et al., 2022)) highlights the importance of rigorous experimental protocols. This discrepancy underscores how easily performance can be overstated in clinical AI applications when methodological rigour is not maintained. Compared to the recurrent models in (Shenoy et al., 2021), our CNN approach shows competitive results, especially for the pressure representation, although slightly lower in F1-score. This suggests that CNNs, when trained on well-represented and properly split data, remain a viable and scalable solution for this problem, while also being simpler to train and deploy than RNN-based models.

Beyond classification performance, this work contributes a broader perspective on the utility of figure-copying tasks in neurodegenerative disease assessment. Simple drawing tasks, such as spirals and lines, have long been recognised as clinically useful for evaluating motor impairments in PD (Alty et al., 2017). Moreover, similar tasks, including clock drawing and wire cube copying, have been effective in detecting cognitive decline and dementia (Costa et al., 2022; Alty et al., 2015). Cognitive and motor impairments have also been jointly investigated using the same dataset and drawing types as the current study. In previous work (Vallejo et al., 2016), clinically-informed features were extracted and selected via a genetic algorithm to estimate MoCA and MDS-PDRS-3 scores using linear models. Unlike that earlier approach, which relied on engineered features, the present study adopts a data-driven method that processes raw drawing signals directly into image-based inputs for CNN training.

Although GAN-based augmentation has been explored in earlier studies (Rodríguez-Martín et al., 2023) for drawing-based PD detection, our work provides several key advantages. Unlike prior work, which focused on synthesising only a limited set of drawing tasks or applied GANs without comparison to baseline augmentation techniques, our study conducts a comprehensive evaluation across three distinct input representations (time, pressure, angle) and compares traditional augmentation with both basic forms of GANs and DCGAN approaches. Moreover, we assess these methods under strict subject-level cross-validation, providing a more realistic estimate of generalisability and robustness. This combination of methodological rigour, representational diversity, and comparative analysis enhances the reproducibility and clinical relevance of our findings.

Despite these contributions, several limitations must be acknowledged. The dataset remains relatively small, imbalanced, and drawn from a single clinical site, limiting generalisability. Additional clinical variables such as medical comorbidities, on-off status, presence of tremor, and anxiety and depression scores (all of which can affect hand dexterity) were not available, hindering deeper analyses. The exclusive use of dominant-hand drawings may have excluded relevant asymmetry information. Additionally, it is worth noting that the PD group had lower cognitive scores, and cognitive decline is known to influence motor performance. In particular, significant cognitive impairment in PD has been associated with increased reliance on visual feedback during upper limb movements, which may in turn affect drawing performance (Cosgrove et al., 2021). Furthermore, generating synthetic samples with GANs and DCGANs proved computationally intensive, often requiring over 30,000 epochs, and occasionally introduced implausible image patterns. Finally, the reliance on specialised digitising tablets may limit broader adoption in less-equipped clinical settings.

Future work should aim to validate these findings in larger and more diverse cohorts, explore online or real-time assessment pipelines, and evaluate the integration of multimodal inputs. Previous work has shown that multimodal approaches combining speech, handwriting, and gait data can improve the classification of neurodegenerative conditions, such as the study of Vásquez-Correa et al. (2018) and Huang et al. (2023). Investigating fusion approaches across time, pressure, and angle signals, or even combining drawing with speech or gait data, may further enhance performance and clinical relevance. The CNN baseline used in this study could be further improved by integrating more efficient deep learning architectures, such as those incorporating transfer learning and attention mechanisms. Combining traditional augmentation with GAN-based techniques may yield more robust and diverse training datasets, while the availability of more data could open the door to applying Dosovitskiy et al. (2020)’s Vision Transformer, potentially improving generalisation and interpretability. Ultimately, our findings support the potential of deep learning applied to drawing tasks as a promising, accessible, and non-invasive tool for early PD screening and monitoring. These results demonstrate that combining diverse input modalities with rigorous cross-validation and systematic augmentation strategies can offer a more realistic benchmark for future clinical AI systems.

## 6. Conclusion

Prompt diagnosis of PD is critical for effective treatment and disease management. This study evaluated the use of deep learning techniques applied to spiral drawing tasks, transforming them into time, pressure, and angle representations. Among these, the pressure-based input combined with traditional geometric augmentation yielded the best performance (80.14% accuracy), supporting its value in discriminating PD from control subjects.

Our findings demonstrate the potential of this approach as a rapid, non-invasive screening tool and highlight the importance of both representation choice and rigorous validation. Future work will focus on optimising the CNN architecture through transfer learning and attention mechanisms, combining augmentation strategies, and exploring transformer-based models as more data becomes available.

## Data Availability

All data produced in the present study are available upon reasonable request to the authors

## Acknowledgements

We would like to acknowledge Qiyue Yang for her initial contributions to the early phases of this project. We were unable to contact her to confirm authorship at the time of submission. Should she wish to be included, we would be happy to revise the author list accordingly.

## Footnotes

1 https://www.tensorflow.org/datasets/catalog/lsun#lsunbedroom

